# Altered plasma, urine, and tissue profiles of sulfatides and sphingomyelins in patients with renal cell carcinoma

**DOI:** 10.1101/2022.01.10.21268223

**Authors:** Robert Jirásko, Jakub Idkowiak, Denise Wolrab, Aleš Kvasnička, David Friedecký, Krzysztof Polański, Hana Študentová, Vladimír Študent, Bohuslav Melichar, Michal Holčapek

## Abstract

Renal cell carcinoma (RCC) represents the most common type of kidney cancer and, despite the progress of surgical and medical management, is associated with high mortality. In this study, we demonstrate that RCC-related processes change body fluids sphingolipid concentrations, which may be used to monitor tumor presence using non-invasive lipid-based blood and urine tests. We investigate 674 plasma, urine, and tissue samples from 369 RCC patients and controls. For the first time, we show the significant concentration changes of low abundant sulfatides in plasma and urine of RCC patients. Elevated concentrations of lactosylsulfatides, decreased concentrations of sphingomyelins with long saturated N-fatty acyls and sulfatides with hydroxylated fatty acyls are the crucial alternations in RCC. These changes are stage-dependent and are more emphasized in late-stage RCC. Similar trends in body fluids and tissues indicate that RCC widely influences lipid metabolism and highlights the potential of lipidomic profiling for cancer detection.

## Introduction

Malignant neoplasms of the urinary system are associated with high rates of mortality and morbidity and constitute a major public health problem. RCC represents the most common type of kidney cancer, and its incidence has increased globally in the last decade^1, 2, 3, 4^. The principal subtype of RCC is clear cell (cc)RCC (>75%), with papillary (pap) RCC (15%) and chromophobe (ch) RCC (<5%) being less common. ccRCC often takes an aggressive behavior, but the tumor biology and outcomes vary substantally^5^.

Early diagnosis improves survival in RCC. Widespread use of abdominal imaging, e.g., ultrasound or computed tomography, has resulted in early detection of RCC in many cases. Renal mass is discovered incidentally during abdominal ultrasonography performed for different complaints, but no laboratory tests distinguish between RCC and benign kidney mass. In many cases, RCC is diagnosed late when metastatic disease is present, and the prognosis is poor^5^. There are no circulating biomarkers for detecting RCC, such as alpha-fetoprotein in hepatocellular carcinoma, carcinoembryonic antigen and carbohydrate antigen (CA) 19-9 in gastrointestinal malignancies, or CA125 and HE4 in ovarian cancer^6^. Therefore, a non-invasive laboratory test to facilitate RCC diagnosis remains an unmet medical need.

Lipidomic analysis may provide information about ongoing processes in the organism. Reliable lipidomics could aid in the detection of cancer^7^, the elucidation of biochemical mechanisms associated with tumor growth^8^, and guide the development of new drugs^9^. For example, sphingomyelins together with ceramides, phospholipids, and glycerolipids showed the potential to detect kidney, breast, pancreatic, and prostate cancer^10, 11^. Sulfatides, a class of low-abundant sphingolipids, also seem to be a good target for such research^12^. The importance of sulfatide metabolism and sulfotransferase genes expression for the morphological differentiation of tumor and metastatic potentials of the colorectal, ovarian, or renal carcinoma was shown in numerous reports^13, 14, 15, 16, 17^. Sulfatides participate in the adhesion to functional proteins, the maintenance and modification of ion channels, and the induction of cell differentiation.

In our previous work, we compared 157 tumor and autologous nonmalignant tissues from 80 patients with RCC^18^, and we reported decreased concentrations of monohexosyl sulfatides (SHexCer) with additional hydroxyl in the ceramide part in the tumor tissue compared to the non-neoplastic autologous tissue. These observations are consistent with the results reported by Kim et al.^13^. We also found elevated levels of sulfatides composed of two hexosyl units (SHex2Cer) in tumor tissues compared to adjacent nonneoplastic tissues. These results are in agreement with previously described enhanced cerebroside sulfotransferase activity in RCC cells^12, 16, 19, 20^. Recent reports also indicate that lactosylsulfatides are involved in cellular adhesion and are suspected to play an important role in the metastasis of hepatocellular carcinoma^21^.

Significantly altered sulfatide profiles are usually observed in tumor tissues or cells because of the high local concentrations of studied lipids. However, only a few studies have been published on sulfatide analysis in cells and tissues of RCC patients ^13, 18^. To the best of our knowledge, no work on the analysis of sulfatides in body fluids of RCC patients has been reported to date in the literature.

Here we compare sulfatide and sphingomyelin profiles of plasma and urine samples taken from RCC patients and controls. We also investigate whether the changes in sulfatide and sphingomyelin profiles of tumor tissues are reflected in body fluids.

## Results

### Study design and analytical validation

A significant part of the cancer samples were ccRCC (128) taken from patients with different tumor stage, including patients with metastatic disease. Patients with other RCC subtypes were also studied, including 15 cases of papRCC, 4 cases of chRCC, 3 cases of multilocular cystic renal cell carcinoma (mcRCC), and 5 cases of patients with other kidney tumors, such as adult cystic nephroma, collecting duct carcinoma, oncocytic adenoma, eosinophilic granular renal cell tumor, and Wilms tumor (see Fig. 1 and Supplementary Data 1-9).

**Fig. 1.**
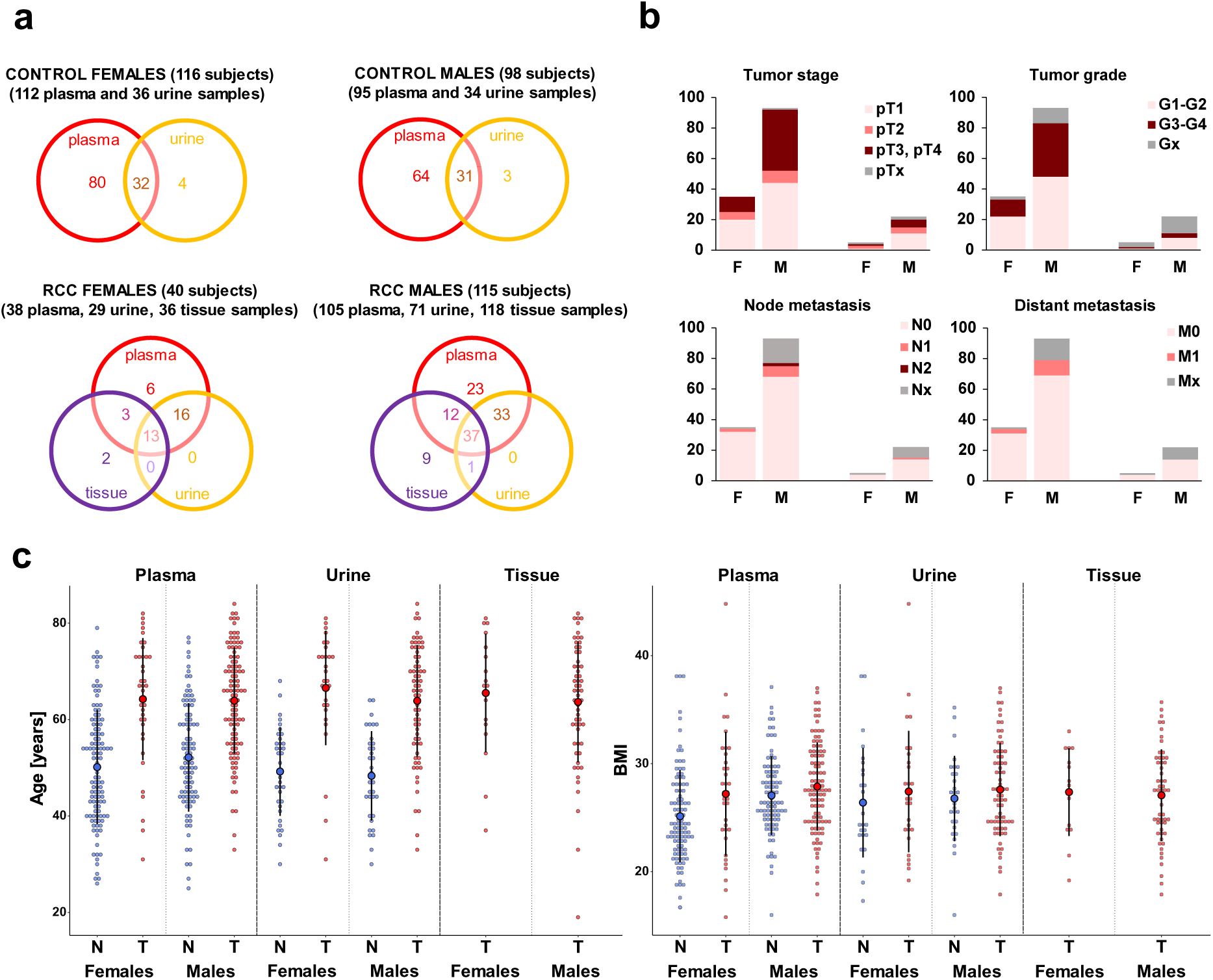
Overview of the number of samples and clinical information. **a** Venn diagrams show the number of individual plasma, urine, and tissue samples obtained from control volunteers and kidney cancer patients (separately illustrated for females and males). **b** Stacked bar charts reflect the TNM tumor classification of cancer patients. Graphs show the number of patients grouped by individual tumor stages, tumor grades, and type of node and distant metastases, separately shown for males (M) and females (F) (see Supplementary Data 1-9 for more details). **c** Dot plots illustrate the age and BMI distribution of control volunteers (N, blue dots) and cancer patients (T, red dots) in the case of plasma, urine, and tissue samples.

In total, we have collected and further investigated 143 plasma, 100 urine, and 154 tissue samples (tumor and non-affected adjacent tissue parts from 77 patients) from 155 kidney cancer patients together with 207 plasma and 70 urine samples from 214 control volunteers (in the following text referred to as controls). The distribution of the sample types from cancer patients or controls is illustrated in Fig. 1a. Both types of body fluids (plasma and urine samples) were investigated for 99 patients with RCC, among whom tissue samples were also evaluated in 50 cases. The higher number of male cancer patient samples in this study reflects that RCC patients are more often males and typically have larger tumors with higher stage and grade than females.

The analytical methodology was validated to ensure high data quality (see Methods). The validation results show that the methodology can be used to quantify sulfatides, sphingomyelins, and sterol sulfates. The measurement of NIST plasma shows comparable results to previously published data (Supplementary Fig. 1)^22, 23, 24^ QC samples were included in the sequence measurements to control instrument performance and data acquisition quality. We have established inclusion criteria for measured lipids before constructing statistical models or calculating relative concentrations (Methods, Data Processing). Data distribution was checked for all quantified lipids within the studied groups, resulting in the application of nonparametric tests.

### Alternations in the plasma lipid profiles of cancer patients

In the first step, we compared the relative concentrations of 33 lipids (Supplementary Data 4) measured in plasma from 143 kidney cancer patients and 207 controls. We observed a partial group separation in the PCA score plot in Fig. 2a, indicating differences between the lipid profiles of cancer cases and controls. A cluster of quality control samples (green dots) shows good method stability during the measurements. Although cancer patients and controls are not age-balanced (Fig. 1c), both groups contain enough samples to build age-matched models to exclude possible bias based on the age, as presented in Supplementary Fig. 2. We also observed the separation of cases and controls in the PCA score plot in this case. We selected the four most statistically significant lipids for both studied lipid classes (SHexCer and SM) based on the fold change, effect size, and false discovery rate, as demonstrated by the volcano plot in Fig. 2b. These lipids were used to generate a circular dendrogram (Fig. 2c), which also shows a good classification of controls and cancer cases.

**Fig. 2.**
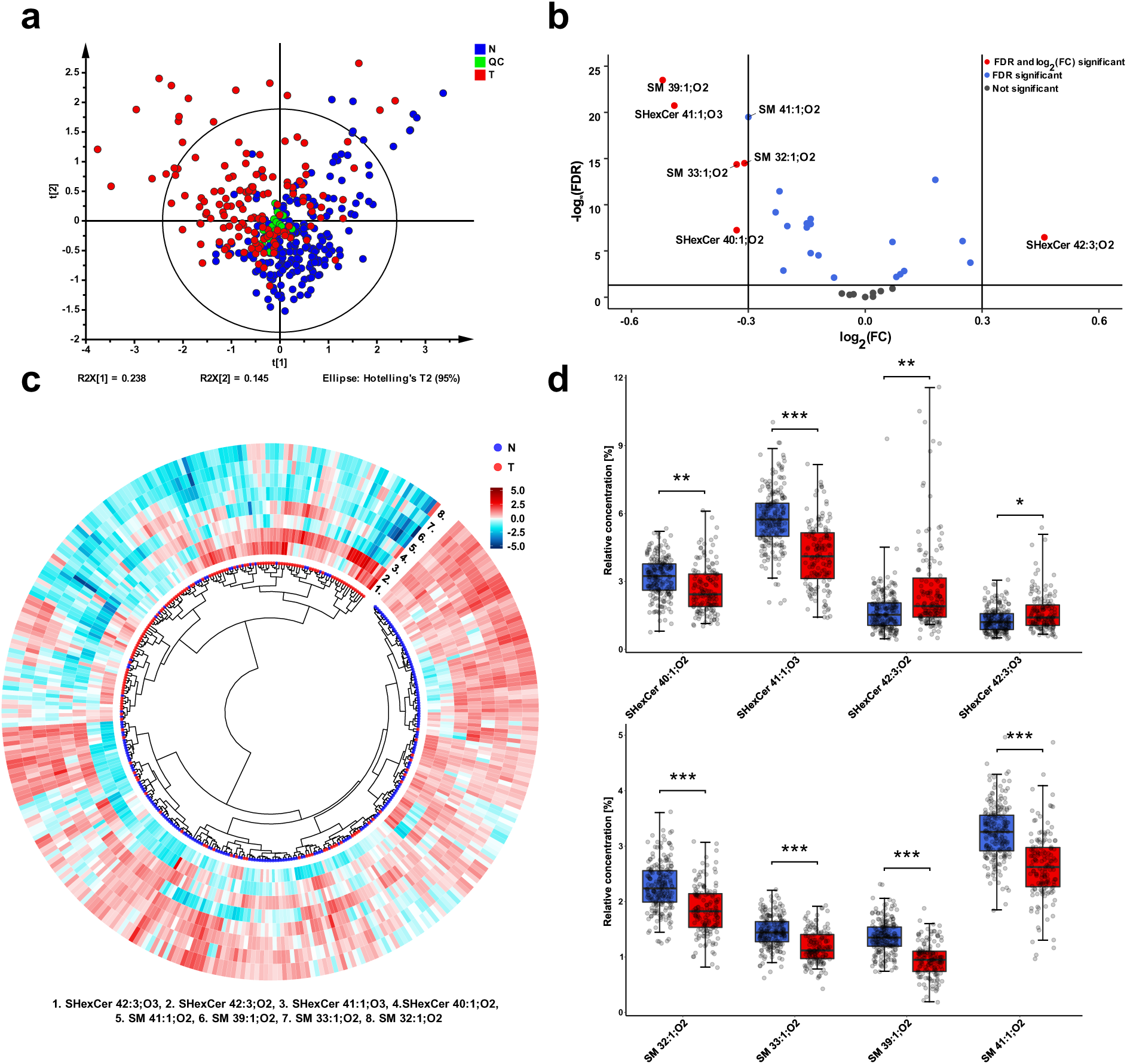
Overview of statistical analysis based on the evaluation of relative concentrations of 33 lipids in plasma samples of RCC cancer patients (T) and controls (N). **a** PCA-X score plot includes 207 controls (blue), 143 cancer patients (red), and 18 QC (green). Data were log-transformed and Pareto-scaled before the PCA analysis. **b** Volcano plot reflects the statistically most significant lipid species with respect to the highest fold change (robust Hodges-Lehmann type fold change estimator) and false discovery rate (FDR) from the Mann-Whitney U test. **c** Circular dendrogram with heat map calculated based on the relative concentrations of 4 most significant SM species and 4 most significant SHexCer species demonstrating the classification of controls from RCC cancer cases. Data were log-transformed and Pareto-scaled, then the Euclidean distances were calculated, and Ward’s clustering method was selected. The heatmap was generated using z-score scaling. **c** Box plots of the relative concentrations of four most significant SHexCer and four most significant SM in plasma samples for controls and patients with RCC. Box plots were adjusted for skewed distributions according Hubert & Vandervieren^55^ . The importance marked above box plots includes fold change, effect size, and FDR from the Mann-Whitney U test: **** - very large, *** - large, ** - medium, * - small significance (more explanation in the Suppl. material 4 - plasma).

Statistical analysis results for all 33 lipids are listed in Supplementary Data 10. In general, most of the studied lipids are downregulated in cancer patients compared to controls, as demonstrated by the volcano plot (Fig. 2b) and box plots (Fig. 2d). The decreased concentrations of SM 41:1, SM 40:1;O2, SM 39:1, SM 38:1, SM 33:1, and SM 32:1 are the most significant changes among sphingomyelins. Sulfatides containing one double bond and mostly with additional hydroxylation (abbreviated as SHexCer XX:1;O3), such as SHexCer 40:1;O3, SHexCer 41:1;O3, SHexCer 42:1;O3 together with SHexCer 40:1;O2, are another significant downregulations in cancer patients. In turn, sulfatides with more double bonds, such as SHexCer 42:3;O2, SHexCer 42:3;O3, and SHexCer 42:2;O2, are upregulated in cancer patients.

The comparison of cancer patients and controls, based on molar concentrations of individual lipids, is listed in Supplementary Data 11 and Supplementary Fig. 3. Downregulation of sphingomyelins containing one double bond, i.e., SM 32:1;O2, SM 33:1;O2, SM 39:1;O2, SM 40:1;O2, SM 41:1;O2, and sulfatides containing one double bond and additional hydroxylation are the most significant differences between RCC patients and controls, similarly, as for the relative concentrations. Only sulfatides with one hexosyl unit were reliably quantified in plasma samples. Such sulfatides are synthesized from glucosylceramides. More complex sulfatides with more than one hexosyl unit are obtained from lactosylceramide. Unfortunately, these lipids were usually below LOQ in plasma, except for SHex2Cer 42:2;O2 (higher concentrations than LOQ were observed for 24 cancer patients and 2 controls). In the case of lipids, which were detected in less than 50% of RCC cases or controls, we decided to use the present/absent approach (Supplementary Data 12). In this way, we investigated if a lipid occurs more often in the plasma of controls or cancer patients (N or T). Results of the analysis are presented using radar charts (Supplementary Fig. 4a). We observe that very low abundant sphingomyelins and sulfatides with one double bond and additional hydroxylation are usually more often present in the plasma of controls than patients. In turn, SHex2Cer 42:2;O2 is present more often in the plasma of RCC patients. Elevated SHex2Cer 42:2;O2 was also found by us previously in tumor tissue^18^. In general, we observe substantial differences between plasma sulfatide and sphingomyelin profiles in RCC patients compared to controls.

### Alternations in the urine lipid profiles of cancer patients

After the data filtration described in Methods, 76 samples taken from RCC patients and another 67 from controls were used to perform the statistical comparison for urine (Supplementary Data 13).

We quantified 18 SHexCer and 4 SHex2Cer in urine. No SM were detected because the concentrations were below the limit of detection, but 11 sterol sulfates (StS) (Supplementary Table 1) were quantified together with sulfatides to provide additional information (Supplementary Data 5). We again observe a partial separation of controls from cancer cases in the PCA score plot (Fig. 3a). A circular dendrogram with a heat map was prepared for 8 selected sulfatides (Fig. 3b), and it also shows some differences between RCC and control samples. In the urine of patients with RCC, concentrations of lactosylsulfatides are elevated compared to control samples. The differences are presented in the heatmap surrounding the dendrogram (Fig. 3b), volcano plot (Fig. 3c), and in the form of box plots (Fig. 3d). Higher levels of SHex2Cer 34:1;O2, SHex2Cer 40:1;O2, SHex2Cer 42:1;O2, and SHex2Cer 42:2;O2 are most significant upregulations. Sulfatides with an additional hydroxyl group in the ceramide backbone (SHexCer XX:XX;O3 or SHexCer XX:XX;O4) show the opposite dysregulation, similarly as in the case of plasma samples. These results might indicate that the synthesis of hydroxylated sulfatides is suppressed in cancer.

**Fig. 3.**
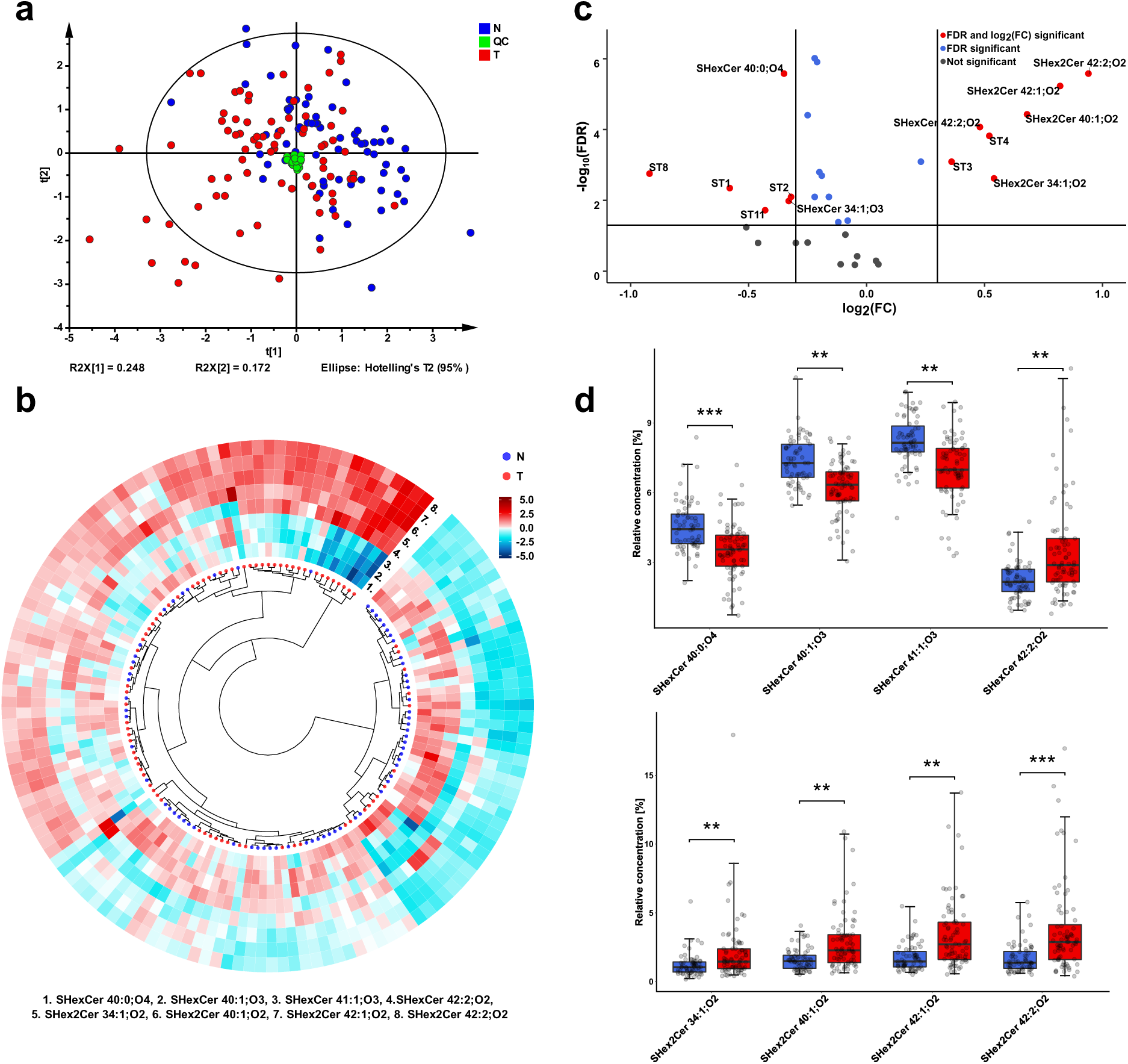
Overview of the statistical analysis based on the evaluation of relative concentrations of 33 lipids in urine samples of RCC cancer patients (T) and controls (N). **a** PCA-X score plot includes 67 controls (blue), 76 cancer patients (red), and 18 QC (green). Data were log-transformed and Pareto-scaled before the PCA analysis. **b** Circular dendrogram with heat map calculated based on the relative concentrations of 4 most significant SHex2Cer and 4 most significant SHexCer species that demonstrates the classification of RCC cancer patients and controls. Data were log-transformed and Pareto-scaled, subsequently the Euclidean distances were calculated, and Ward’s clustering method was selected. The heatmap was generated using z-score scaling. **c** Volcano plot reflects the statistically most significant lipid species with respect to the highest fold change (robust Hodges-Lehmann type fold change estimator) and false discovery rate (FDR) from the Mann-Whitney U test. **d** Box plots of relative concentrations of 4 most significant SHex2Cer and 4 most significant SHexCer for controls and patients with RCC cancer. Box plots were adjusted for skewed distributions according Hubert & Vandervieren^55^. The importance marked above box plots includes fold change, effect size, and FDR from the Mann-Whitney U test: **** - very large, *** - large, ** - medium, * - small significance (more explanation in the Suppl. material 13 - urine).

The present/absent analysis results also show that hydroxylated sulfatides occur more often in the urine of controls than RCC cases (Supplementary Fig. 4b, Supplementary Data 14). Another difference between the urine samples of RCC patients and the controls is observed in the case of sterol sulfates. In patients with RCC, we observe upregulation of StS 3 and StS 4 and downregulation of StS 8 and StS 11. In summary, we also found differences between the urine lipid profiles of RCC cases and controls

### Different sulfatide and sphingomyelin levels in patients with advanced tumor stage and grade

To answer whether the lipid changes observed in both biological fluids are associated with tumor stage and grade, we divided the plasma and urine sample sets into controls, and patients with early (T1-T2) and more advanced tumors (T3-T4). Results of statistical tests are presented in Supplementary Data 15 and 16 for plasma samples and Supplementary Data 17 and 18 for urine samples, respectively.

The box plots in Fig. 4 and Supplementary Fig. 6 illustrate a gradual drop in concentrations of O3 and O4 hydroxylated sulfatides SHexCer XX:XX;O3 or SHexCer XX:XX;O4 in plasma and urine samples with higher stage and grade. In contrast, a gradual increase in concentrations of lactosylsulfatides SHex2Cer XX:XX;O2 and monohexosylated sulfatides SHexCer 42:2;O2 and SHexCer 42:3;O2 was observed with increasing cancer stage and grade. Based on these results, we suspect that changes in the profile of sulfatides might be related to tumor progression, and we hypothesize that cancer can affect sulfatide metabolism. Similarly, lower levels of SM with longer fatty acyls, i.e., SM 39:1, SM 40:1, and SM 41:1 were found in patients with more advanced tumor stage or grade. These findings show that in RCC, plasma and urine lipid profiles differ slightly between patients, depending on how advanced the tumor is.

**Fig. 4.**
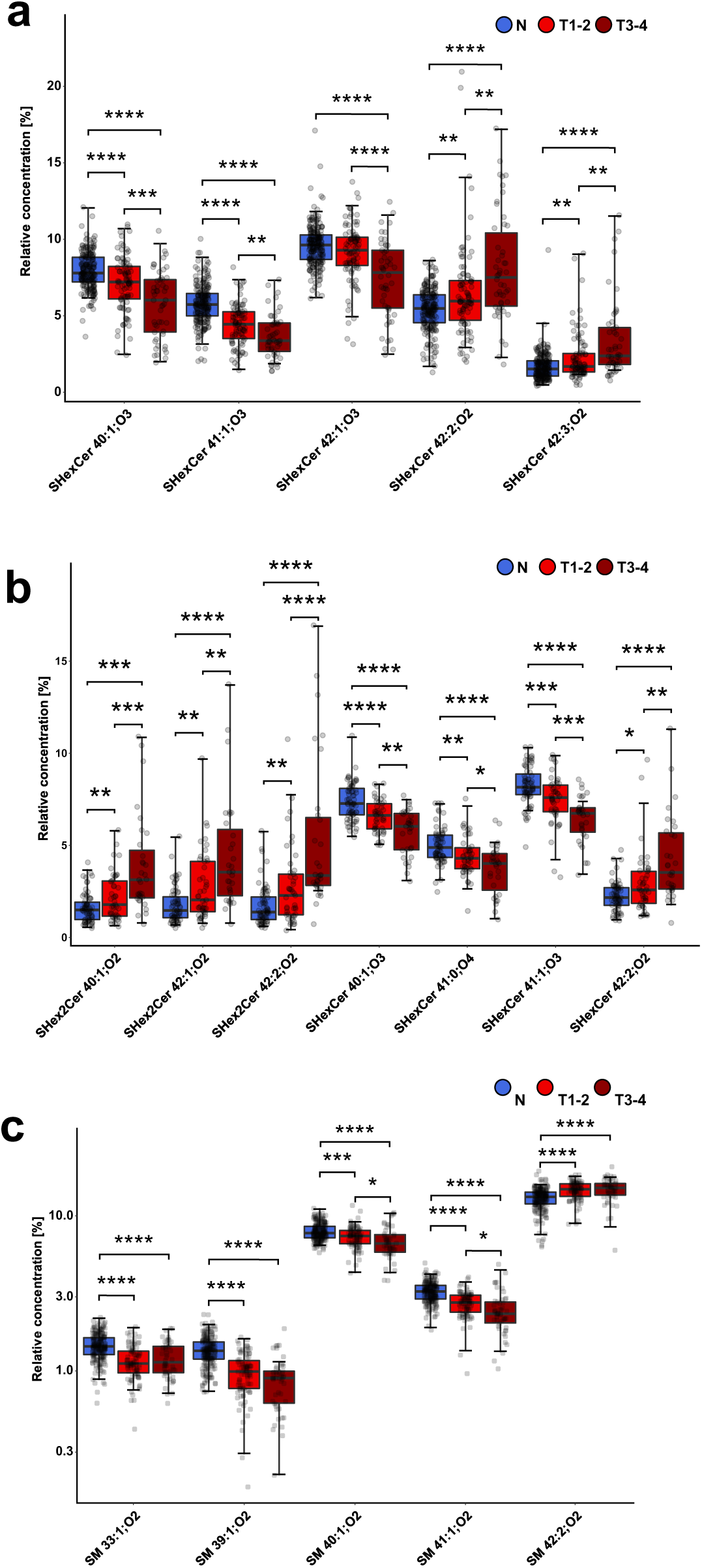
Gradual dysregulation in cancer samples with increased tumor stage. Box plots of the most statistically significant lipids for controls, patients with tumor stages T1-2, and T3-4: **a** sulfatides in plasma samples, **b** sulfatides in urine samples, and **c** SM in plasma samples. Box plots were adjusted for skewed distributions according Hubert & Vandervieren^55^ . The importance marked above box plots indicates results of the Conover posthoc test (FDR): 0.05-0.01 - *, 0.01-0.001 - **, 0.001-0.0001 - ***, 0.0001 and lower - ****.

### Observed sulfatide and sphingomyelin dysregulations in RCC patients are similar in body fluids and tumor tissues

To support the hypothesis that dysregulation observed in plasma and urine can result from cancer progression, we further investigated differences between lipid profiles of tumor and autologous nonneoplastic tissues of 77 patients (Supplementary Data 6). Results of statistical tests for 44 quantified lipids, including 20 SHexCer, 12 SHex2Cer, and 12 SM species, are listed in Supplementary Data 19. The separation of both sample groups was observed in the PCA score plot (Fig. 5a).

**Fig. 5.**
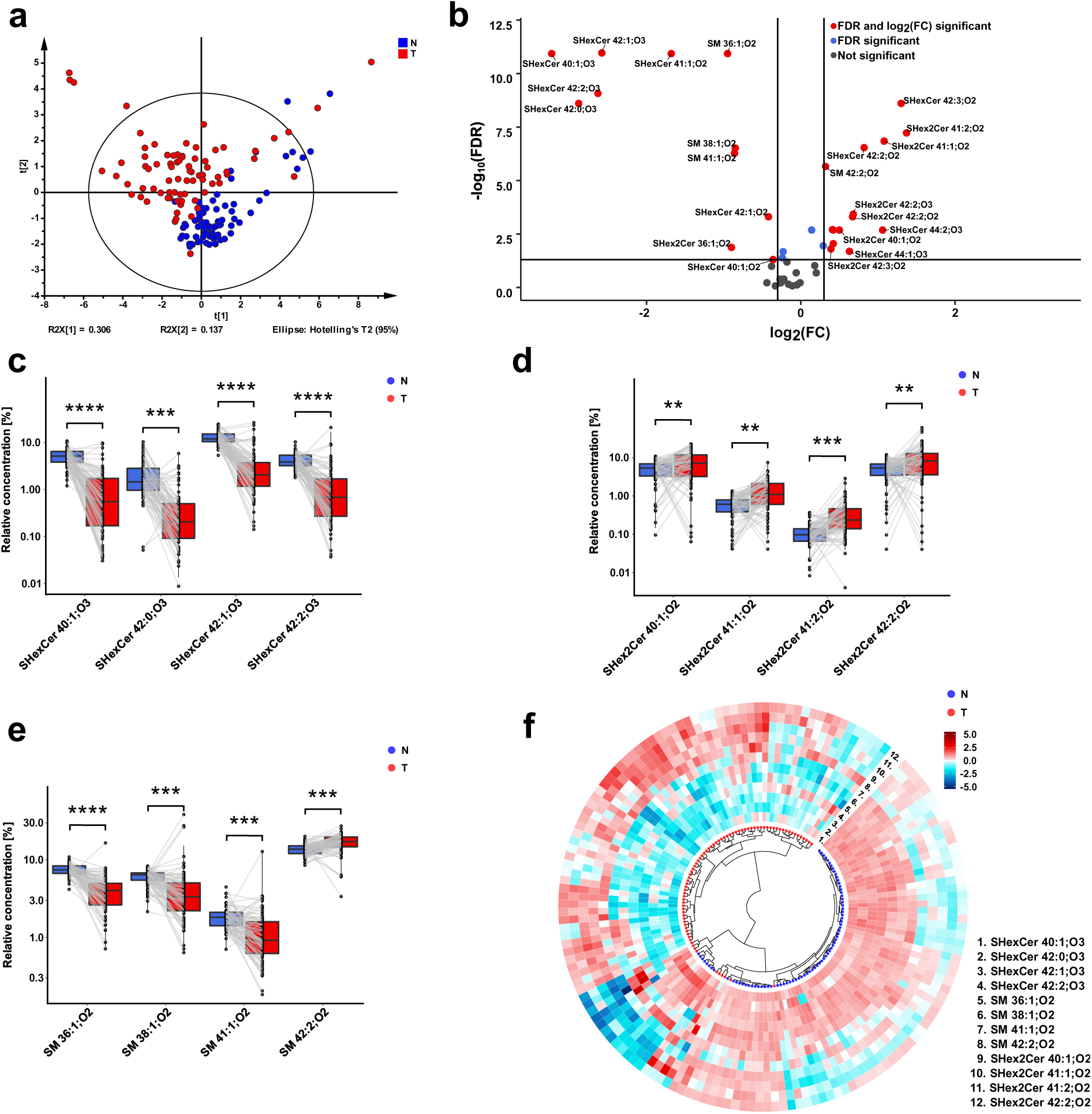
Overview of statistical analysis based on the evaluation of relative concentrations of 44 lipids in nonneoplastic (N) and tumor (T) tissues of 77 patients. **a** PCA-X score plot includes all 144 studied tissue samples. Data were log-transformed and Pareto-scaled before the PCA analysis. **b** Volcano plot reflects statistically most significant lipid species with respect to highest fold change (robust Hodges-Lehmann type fold change estimator) and false discovery rate (FDR) from the Wilcoxon signed-rank test for paired samples. **c-e** Box plots of concentrations of 4 most significant SHexCer, 4 most significant SHex2Cer, and 4 most significant SM in tumor tissue and nonneoplastic tissue samples of RCC patients. Box plots were adjusted for skewed distributions according Hubert & Vandervieren^55^. The importance marked above box plots includes fold change, effect size, and FDR from the Wilcoxon signed-rank test: **** - very large, *** - large, ** - medium, * - small significance (more explanation in the Suppl. material 19 - tissues). **f** Circular dendrogram with heat map calculated based on the concentrations of 4 most significant SM, 4 most significant SHexCer, and 4 most significant SHex2Cer species demonstrates classification of tumor and nonneoplastic tissue samples of RCC patients. Data were log-transformed and Pareto-scaled, then the Euclidean distances were calculated, and Ward’s clustering method was selected. The heatmap was generated using z-score scaling.

When we compared lipid profiles of two tissue types, we found similar changes to these observed in body fluids. Namely, in the tumor tissues, hydroxylated sulfatides and SM with one double bond, such as SM 41:1;O2 are downregulated. In contrast, elevated SHex2Cer species, SHexCer 42:2;O2, SHexCer 42:3;O2 and SM 42:2;O2 are observed in tumor tissues as illustrated in the volcano plot in Fig. 5b or the paired box plots in Fig. 5c-e. The alterations in SHex2Cer, SHexCer, and SM are also presented in the circular dendrogram’s heat map (Fig. 5f). In the dendrogram, nonneoplastic and tumor tissue samples are well-separated from each other.

The results from the present/absent analysis for lipids, which did not fulfill inclusion criterion 1, in the case of tissue samples, are summarized in Supplementary Data 20. Because both tissue parts were taken from one subject with RCC, we considered the following cases: lipid present (or absent) in nonneoplastic/tumor tissue at once, i.e., (1,1) or (0,0), present only in nonneoplastic tissue (1,0) or only in tumor tissue (0,1). The McNemar’s test was applied for the presence/absence analysis in this case. Similarly to plasma and urine results, we found that hydroxylated sulfatides and SM are present more often in nonneoplastic parts but absent in the tumor tissue, i.e., usually more (1,0) pairs are found than (0,1) for SM and hydroxylated sulfatides.

Then, to see the whole picture, we compared all alterations in lipid profiles found in RCC vs. control samples across all three types of studied materials, as illustrated in network visualization (Fig. 6).

**Fig. 6.**
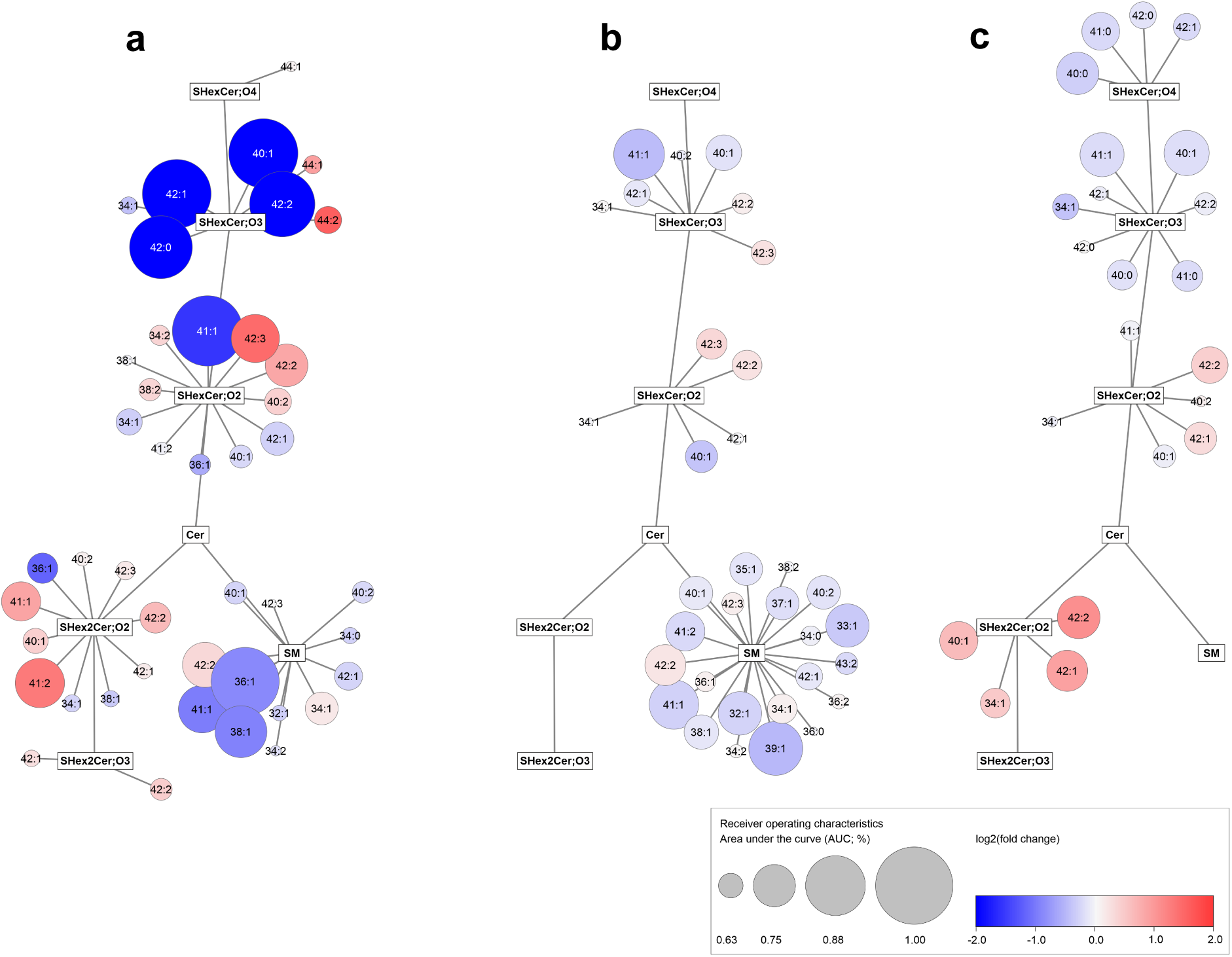
Network visualization of dysregulated lipids in RCC across all three types of samples. Graphs show simplified metabolism of sphingolipids **a** tissue, **b** plasma, and **c** urine samples; the Cytoscape software (http://www.cytoscape.org) was used to generate the networks. Circles represent the detected lipid species, where the circle size expresses the significance according to AUC values from ROC analyses listed in Supplementary Data 27, while the color saturation defines the degree of up/downregulation (red/blue).

Similar dysregulation trends in all RCC samples might indicate that lipid alternations originated in tumor tissue are reflected in body fluids studied. However, no gradual drop with increasing tumor stage (Supplementary Data 21 and 22) or grade (Supplementary Data 23 and 24) is observed in the case of tissue samples, as presented in Supplementary Figures 7 and 8. Patients with advanced tumor are at increased risk of developing metastases, and the advanced disease might result in stronger differences between their lipid profiles and controls. We compared lipid concentrations in body fluids (plasma and urine) of patients with cancer stage T3 - with and without metastases (Supplementary Data 25 and 26, and Supplementary Fig. 9). Increased concentrations of SHex2Cer species and SHexCer 42:2;O2, are observed in the urine of patients with metastases, compared to patients with no metastases. In turn, hydroxylated sulfatides in the urine and plasma and SM in plasma are lower in patients with metastases. It supports the hypothesis that cancer metastases are associated with higher degree of lipid dysregulations.

### Potential of studied lipids for clinical cancer detection

To check the potential of the lipids studied for clinical cancer detection, we divided plasma samples into a training and testing cohort, and about 25% of all samples were allocated to the testing set.

Relative concentration of 33 lipids, presence/absence data for 13 lipids, gender, and BMI were used to build the classification model. We selected logistic regression with ridge penalty. The influence of clinical parameters on lipid concentrations was analyzed before the model was built, including gender, age, and BMI. All subjects were split according to gender and then – into controls, RCC patients with tumor stage T1-2, and patients with tumor stage T3-4. After splitting the data according to gender, we observed that the respective medians in box plots are slightly shifted, and gender separation might improve the differentiation of cases from controls (Supplementary Figure 10). Spearman correlations were calculated between lipid concentrations and age (Supplementary Table 2) and lipid concentrations and BMI (Supplementary Table 3). Most of the obtained correlations indicate a relatively weak association between age and lipid concentrations or BMI and lipid concentrations and are not statistically significant. Moderate positive correlations between BMI and SM 36:0;O2, SM 36:1;O2, and SM 36:2;O2 levels, and moderate negative correlations between BMI and SHexCer 42:1;O2, SHexCer 42:2;O2, SHexCer 42:3;O2 levels may indicate alterations in the metabolism of C18 and C24-based sphingolipids in obesity. Besides, stronger correlations between lipids and BMI for females with T3-4 cancer stages may be associated with a low number of samples available for calculations. Finally, gender and BMI were included in the model, but we resigned from the age because it was not well-balanced between controls and RCC patients (Fig.1). The ability of the model to distinguish controls from patients in the case of plasma samples is illustrated in Fig. 7a. The value of the area under the receiver operating characteristic curve (AUC) was 0.973 (95% confidence interval range from 0.940 to 0.995) for the testing set and 0.940 for the training set (95% confidence intervals range from 0.909 to 0.965). The sensitivity, specificity, and accuracy for the classification of training and testing samples are presented in the form of a bar chart (Fig.7b).

**Fig. 7.**
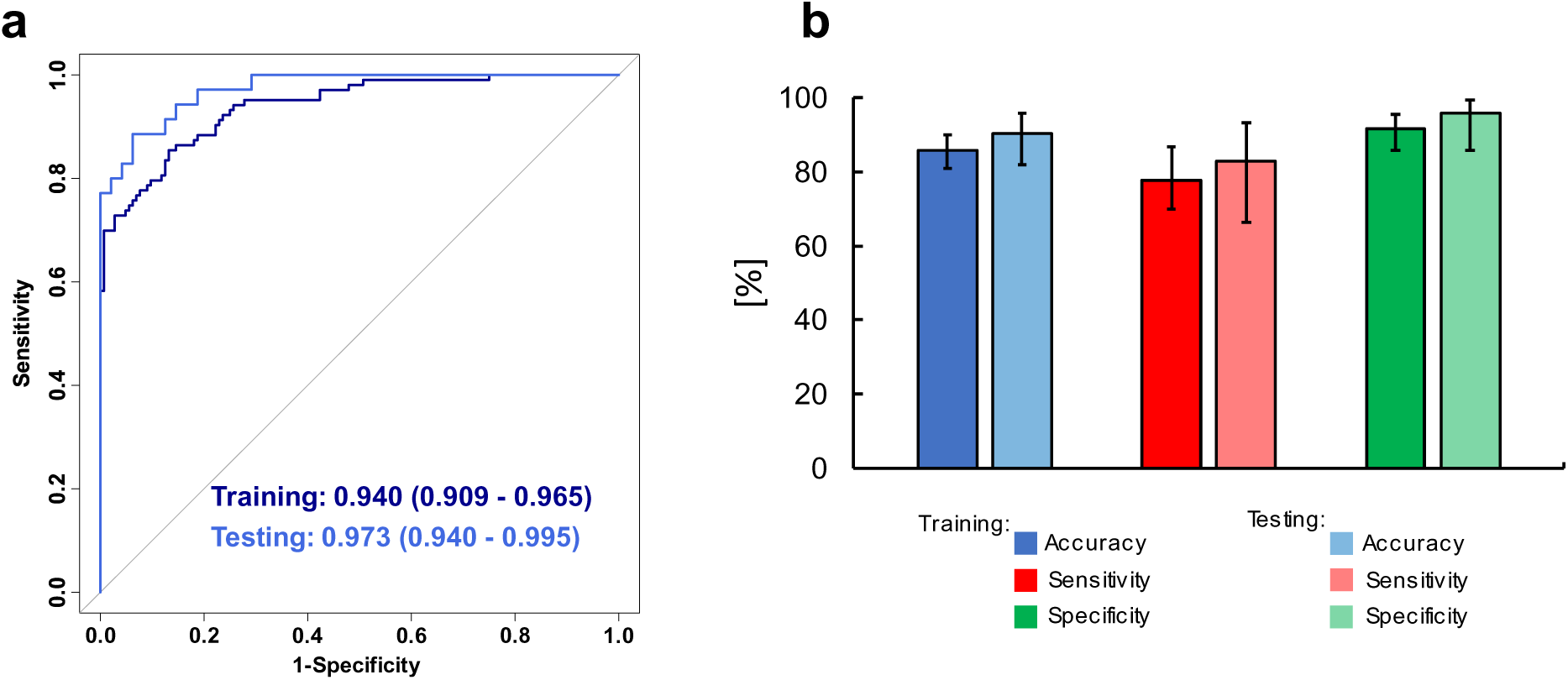
Potential of the studied lipids in case of clinical cancer detection: **a** ROC curves based on logistic regression with ridge penalty for the training and testing sets. Data from 207 controls and 143 RCC patients were divided into training (75%) and testing set (25%). **b** The obtained sensitivity (red) specificity (green), and accuracy (blue) values in percentage for the training and validation set. The error bars represent the 95% confidence intervals for accuracy, sensitivity, and specificity.

The clinical potential of studied sulfatides and sphingomyelins in all studied materials is further evident in the network visualization of dysregulated lipids (Fig. 6). The circle size expresses the significance according to AUC values from ROC analyses listed in Supplementary Data 27.

## Discussion

Metabolic reprogramming is a hallmark of cancer and plays an important role in malignant transformation and tumor progression, shapes tumor microenvironment, and the phenotype of cancer cells, further contributing to tumorigenesis^25^. Sphingolipid metabolism shows an imbalance in many cancers, which is consistent with the involvement of these lipid classes in basic cellular mechanisms, including proliferation, migration, and senescence^26^. Lipids belonging to sphingolipids, such as ceramides, glucosyl-/galactosyl- and lactosyl-ceramides, and sphingomyelins, have been proposed several times as potential biomarkers for the early detection of kidney, breast, prostate, or pancreatic cancer^7, 10, 11, 27^.

Recently, we have reported significant concentration changes of sulfatides in RCC tumor tissues compared to the nonneoplastic tissue part. One of the most important alternations, downregulation of hydroxylated monohexosylsulfatides (SHexCer XX:XX;O3 and SHexCer XX:XX;O4), is also observed in the all three types of biological matrices in the present study. The structure of the sphingoid base in sulfatides is generally predominantly represented by sphingosine C18, while the N-acyl chain exhibits a greater degree of structural variability in both the length and in the range of metabolic modifications, such as desaturation or hydroxylation^28^. The constituents of one of the typical modified N-acyl types are 2-hydroxy fatty acids (hFA), and from them, the corresponding hFA-sphingolipids are formed. The precursors of hFA-sphingolipids, hydroxy fatty acids (hFAs), are enzymatically formed from fatty acids catalyzed by fatty acid 2-hydroxylase (FA2H, EC 1.14.18.6)^29^. The decrease in FA2H expression and associated reduced levels of hFA-sphingolipids have been described in aggressive and treatment-resistant tumors. Altered FA2H expression may be associated with resistance to treatment^30^ and the progression of tumor growth, for example, in lung^31^, ovarian^32^, colorectal^33^, and breast cancer^34^. In turn, increased expression of FA2H and higher levels of hFA-sphingolipids have been described in chemosensitive tumors and patients with a better prognosis^30, 32^. Therefore, we hypothesize that determining hydroxylated sulfatides in plasma and urine might have a diagnostic value in RCC.

In tumor tissues and urine from RCC patients, elevated sulfatide content, especially dihexosyl-sulfatides (SHex2Cer XX:XX;O2), is another crucial dysregulation and might be related to the metabolism of glutamine. Increased glutamine uptake and its utilization in the TCA cycle or as a nitrogen and carbon donor have been observed in many cancers^35^, specifically in RCC^36, 37^. The dysregulation of glutamine metabolism increases the secretion of ammonium ions, and sulfatides, as anionic molecules, carry a negative charge that can balance the positively charged ammonium ions. This mechanism was already proposed in chronic metabolic acidosis, where renal sulfatides serve the same purpose^38, 39^.

Concerning sterol sulfates in urine samples, the upregulation of StS 3 and StS 4 (Fig. 3c and Supplementary Fig. 5) corresponding to elemental compositions of cortisol metabolites (Supplementary Table 1) indicates increased stress in patients with RCC^40^. In contrast, the downregulation is observed for sterols StS 8 and StS 11 with the elemental composition corresponding to lithocholic acid metabolites. As lithocholic acid has recently been shown to induce apoptosis within several tumor cell lines^41^, its suppression may result from cancer metabolic reprogramming.

Another class of sphingolipids investigated in our work is SM that are reduced in the tumor tissue as well as in the plasma of RCC patients. For example, SM 41:1 and SM 42:1 are downregulated in both sample matrices, but lower levels of multiple other SM are observed in plasma rather than in the tumor tissue. Reduced levels of SM in RCC have been documented before ^10, 27^. In contrast, higher levels of SM were associated with the antitumor effect of 2-hydroxyoleic acid (2OHOA)^42^. However, the exact mechanism of action has not yet been elucidated and probably it is not related to direct activation of sphingomyelin synthase (SMS, EC 2.7.8.27)^43^. Furthermore, SM-enriched cells are more sensitive to apoptosis^44^. This biochemical alteration may be explained by the upregulation of SM to ceramide conversion pathway in tumor tissue via SMS. SMS, which exists in isozyme variants SMS1 and SMS2, is the only enzyme that produces SM in humans by transferring of phosphocholine from phosphatidylcholine to ceramide^45^. The role of SMS in cancer is complex, shows discrepancies, and is widely debated, for example, because expression of SMS1 was found to be reduced in lung cancer, but not significantly altered in esophageal cancer^46^. Additionally, decreased expression of SMS1 has been associated with sphingolipid reprogramming and a worse prognosis in advanced melanoma patients^47^. On the other hand, increased SMS2 expression has been observed in patients with metastatic breast cancer^48^, and SMS2 deficiency led to the inhibition of colon cancer progression in murine model^49^. These findings suggest that SM levels and alteration of the synthetic and degradative pathways play an essential role in cancer. Reduced SM levels observed in RCC samples may promote cancer cell survival. However, the mechanism has not yet been deciphered, mainly due to the complexity and different behavior in various types of tumors.

Sphingolipids have been the focus of many previous studies^7, 10^ looking for biomarkers allowing early kidney cancer diagnosis. However, these studies did not include multi-tissue/plasma/urine comparison of the concentrations of these sphingolipids, and selected biomarkers were often not fully quantified by validated methods. Therefore, these were the key points of our study. Here, we describe the dysregulation of specific sphingolipids and for the first time show significant concentration changes of low abundant sulfatides in the body fluids of RCC patients based on the comparison of blood and urine samples from 358 RCC patients and controls. Compared to previously published reports, we confronted alterations observed in lipid profiles of tissues and biological fluids to analyze how widely cancer-associated processes influence sphingolipid levels. We found that sphingolipids with the same structural elements show a similar dysregulation in all three types of investigated materials. Increased dysregulations in more advanced tumor stage and grade may be used to predict various pathological states of RCC. This suggests the potential application of sphingomyelins and sulfatides for the early detection, assessment of tumor progression, and treatment monitoring of RCC in plasma or urine.

## Methods

### Sample collection

Plasma and urine samples of kidney cancer patients and healthy controls were obtained at the Department of Oncology, Faculty of Medicine and Dentistry, Palacký University and University Hospital Olomouc, Czech Republic (Supplementary Data 4 and 5 for more details). 154 tissue samples, including 77 kidney tumor parts and 77 adjacent nonmalignant kidney tissue parts from 77 RCC patients (Supplementary Data 6 for more details), were obtained from the Department of Urology, Palacký University, Faculty of Medicine and Dentistry and University Hospital, Olomouc, Czech Republic^18^. Controls are samples obtained from volunteers without cancer records (the only exclusion criterion). For cancer patients, the disease was confirmed by biopsy, surgical resection, or histology. All cancer patients and controls were of Caucasian ethnicity, collected at the same place, and processed the same way. The study was approved by the Ethics Committee of the University Hospital Olomouc and Medical Faculty of Palacký University in accordance with Ethical Principles for Medical Research Involving Human Subjects (Helsinki Declaration). All subjects signed an informed consent. Blood and urine collection was performed after overnight fasting with the recommendation to avoid fat-rich meals and alcohol consumption in the evening before the collection. Blood and urine from cancer patients were obtained before surgical operation and treatment. The samples were stored at − 80 °C, transported from the hospital to the analytical laboratory at -20 °C on dry ice, and subsequently stored again at − 80 °C until the sample processing.

### Sample processing

Solvents used for extraction were purchased from Sigma-Aldrich (St. Louis, MI, USA). For lipid extraction from 25 μL of plasma, a modified Folch method with a chloroform – methanol – water mixture was employed^10^. The samples were spiked before extraction with a mixture of internal standards obtained from Avanti Polar Lipids (Alabaster, AL, USA) to achieve final concentrations of 0.1 nmol of SHexCer d18:1/12:0 and 43.3 nmol of SM d18:1/12:0 per ml of plasma. For urine samples, reversed-phase solid-phase extraction (SPE) was performed. Briefly, 2 ml of human urine together with 3 μl of the mixture of internal standards dissolved in methanol (SHexCer d18:1/12:0 of concentration 1.7 μg/ml and D4 taurocholic acid of concentration 16.7 μg/mL) were loaded into a 200 mg tC18 cartridge (Sep-Pak Vac, 37-55 µm particle size) (Waters, Milford, MA, USA) providing concentrations 0.04 nmol of SHexCer d18:1/12:0 and 0.55 nmol of D4 taurocholic acid per ml of urine. SPE columns were previously primed with 3 mL of methanol followed by 3 ml of water. The columns were washed with 3 mL of water, and the studied lipids were further eluted with 3 ml of methanol. The eluates were collected, then evaporated by a gentle stream of nitrogen, and redissolved in the mixture of 600 µl of methanol before the measurement.

### Mass spectrometry analysis

Mass spectra were measured using an ultrahigh-resolution MALDI LTQ Orbitrap XL mass spectrometer (Thermo Fisher Scientific, Waltham, MA, USA) equipped with a nitrogen UV laser (337 nm, 60 Hz) with a beam diameter of about 80 μm x 100 μm. The LTQ Orbitrap instrument was operated in negative-ion mode over a normal mass range *m/z* 400 – 2000, and the mass resolution was set to R = 100,000 (full width at half maximum definition, at *m/z* 400) for all full scan mass spectrum experiments (an identical setting was used for MALDI-MSI experiments). 9-AA (Sigma-Aldrich, St. Louis, MO, USA) was dissolved in methanol (Sigma-Aldrich) – water mixture (4:1, v/v) to provide a concentration of 5 mg/ml for plasma samples or 10 mg/ml for urine samples and mixed with lipid extracts (1:1, v/v). Deionized water was prepared with a Milli-Q Reference Water Purification System (Molsheim, France). The deposited volume of each sample on the target plate was 1 μL and was followed by dried droplet crystallization. Each sample was deposited on MALDI plate five times and measured in negative polarity mode. The zig-zag sample movement with 250 μm step size was used during the individual data acquisition. The laser energy corresponded to 2.7 μJ and 2 microscans/scan with 2 laser shots per microscan at 36 different positions were accumulated for one measurement to achieve a reproducible signal.

### Data processing

Each measurement was represented by 5 average MALDI-MS spectra (5 repetitions) with thousands of m/z values for each sample. The automatic peak assignment was subsequently performed, and m/z peaks were matched with deprotonated molecules [M-H]^-^ for sulfatides and StS or [M-CH_3_]^-^ for SM from a database created during the identification procedure using the Excel macro script available on figshare (https://figshare.com/s/d01d43e862b7e03adbfe). This peak assignment resulted in the generation of the list of present m/z of the studied lipids with the median intensities and relative standard deviations of repetitions for each measured sample, which was used for further IS (calculation of molar concentrations) or relative normalization (calculation of relative concentrations) and statistical evaluation. Obtained lists of all m/z together with their median intensities for all studied samples are presented in Supplementary Data 1-3. For all three types of samples, we have calculated both relative (Supplementary Data 4-6) and molar concentrations (Supplementary Data 7-9). The large differences in case of normalization to IS (Supplementary Data 7-9) within the urine or tissue samples studied resulted in many outliers, therefore we have finally decided to discuss the results of relative concentrations that allow direct statistical comparison of all three types of samples (plasma, urine, and tissue). Molar concentrations were calculated as follows: the intensity of lipid species was divided by the intensity of a particular IS and multiplied by IS concentration. Zero values were replaced by 80% of the minimum within all samples for each corresponding lipid species for molar concentrations. Only lipids, which met two inclusion criteria, have been further used to calculate relative concentrations. The first criterion (inclusion criterion 1) is that the evaluated lipid species must be present at least in 50% of samples in both groups, i.e., controls x cancer cases, of individual datasets (plasma, urine, and tissue). The second criterion (inclusion criterion 2) is that the coefficient of variation of the raw intensities (Supplementary Data 1-3) of the lipids in the measured QC samples must be less than 35%. In the case of urine samples, we have also excluded 27 selected subjects (24 patients and 3 controls) because more than 50% of the lipids were below LOQ or the IS signal was below LOQ due to the high matrix effect (inclusion criterion 3). For the relative quantitation, the zero values (see Supplementary Data 1-3) were replaced by the value corresponding to the lowest reproducible intensity (8000) for all lipids. Signal intensities of each lipid (feature) were related to the sum of all intensities for all lipid species within a particular class, i.e., SM, sulfatides, and sterol sulfates, and multiplied by 100 for each sample separately to calculate relative % (the percentage abundance of lipid species within individual lipid classes).

### Method validation

All used internal standards (SHexCer 18:1;O2/12:0, SM d18:1;O2/12:0, and taurocholic acid-d4) were tested for their applicability for quantitation of plasma or urine samples. The bioanalytical method validation guidelines^50, 51^ were followed and adapted. Representative samples of controls and cases characterized by different gender, age, and health state, were mixed in equal ratios for the pooled sample preparation - QC. The parameters determined included calibration curve, selectivity, repeatability, extraction recovery, matrix effect, within-run and between-run precision, carry-over effect, and freeze/thaw stability. Aliquots of the pooled sample were either not spiked, spiked before or after extraction with a mixture of IS at a low, medium, or high concentration level, respectively. Medium concentration was identical to the IS concentrations in studied plasma and urine samples of controls and RCC patients. Individual IS concentration in plasma and urine are listed in Supplementary Table 4.

The reproducibility of sample spotting is represented by the coefficients of variation (CV) calculated from five IS intensities of 5 consecutive spots of each sample (Supplementary Tables 5-7). All validation parameters were calculated using Microsoft Excel® 2016 (ver. 2102). The investigated parameters were determined on two independent days (except freeze/thaw stability). The linearity range and the limit of quantitation (LOQ) of the developed MALDI-MS method were determined from a calibration curve based on the extracted pooled sample of plasma or urine spiked with IS mixture of different concentration levels (Supplementary Table 8 and Supplementary Figure 11a-d). Calibration curves were plotted as peak abundances against concentrations of IS individual calibration solutions at 8 concentration levels and fitted by linear regression. The measurement of high number of samples with five repetitions is not possible within one run on one MALDI plate. The washing step of the plate is necessary before the next run, and the carry-over effect should be determined to verify the efficiency of the washing procedure. The carry-over effect was determined from the blank sample, which was spotted on the washed MALDI plate on the same positions as previously measured calibration standards at the upper limit of quantitation. No signal at m/z corresponding applied IS was observed for both sample types. The extraction recovery was determined by calculating the ratio of the IS signal intensity of 6 samples (3 for urine) spiked before and 6 samples (3 for urine) spiked after the extraction at three concentration levels (Supplementary Tables 9, 10, and Supplementary Figure 11e, f). The precision was calculated considering the mean value and standard deviation (Supplementary Table 11). Within-run precision was calculated as the CV of IS intensities (IS spiked before extraction) within three consecutive extractions performed in one day for low, medium (only for plasma), and high concentration levels. The between-run precision was determined based on the measurement of six extracts of the pooled samples at three concentration levels (three extracts were prepared in one day, and another three extracts were prepared in the second day). For evaluating selectivity, matrix effect, and freeze/thaw stability of the plasma samples, plasma extracts of six different individuals were measured (Supplementary Table 6). The selectivity of the tested IS was verified based on the evaluation of the data obtained after measurement of these samples without the addition of IS, where no interference signal was observed at *m/z* corresponding to IS. The matrix effect was calculated as the coefficient of variation of IS intensities within the mass spectra of samples of six different individuals at three concentration levels (Supplementary Tables 12). Freeze/thaw stability was determined individually for each sample of 6 individuals with IS at the medium level, and it was calculated as the coefficient of variation of four median intensities within three freeze/thaw cycles involving the storage of the sample at -20°C overnight and its thawing at room temperature for 1 h (Supplementary Table 13). For urine samples, the selectivity of the tested IS was verified based on the evaluation of data obtained after measurement of pooled plasma and plasma samples of six random individuals (without the addition of IS), where no signal at m/z of IS was observed in all cases. Freeze/thaw stability of urine was verified based on the comparison of IS intensities of the pooled sample with IS at low and high concentration levels after one freeze/thaw cycle involving the storage of the sample at -20°C overnight and its thawing at room temperature for 1 h (Supplementary Table 13). High matrix effect was observed for urine samples as evident from the Supplementary Data 1 (raw intensities) of IS intensities in the studied samples resulting in the decision to normalize the data to the sum of all intensities for all lipid species within a particular class for each sample separately to calculate relative concentrations in %.

### Quality control and NIST plasma measurement

Pooled samples (see above) were used as quality control (QC) to monitor the instrument performance and the quality of data obtained in sequence measurement of plasma and urine samples. At least 3 QC samples were spotted on each MALDI plate at the beginning, middle, and end of the sequence measurement. Supplementary Figures 12 and 13 show the variation in absolute intensities of IS and selected endogenously present lipids within all measured QC samples. The middle line represents average intensity, and the dashed lines represent the coefficient of variation of 20%. The coefficient of variation of the absolute signal, which in some cases exceeds 20%, especially for SHexCer 41:1;O3, is reduced by normalization to IS or sum of intensities of all quantified sulfatides (relative concentrations), which is shown in Supplementary Figures 12d and 13f, where the absolute concentration of selected lipids within QC samples are replaced by molar or relative concentrations. Similarly, for urine samples, the coefficient of variation is reduced in the case of monitoring relative concentrations (Supplementary Figures 13). The clusters of QC samples in the PCA score plots indicated method stability during measurements (Fig. 2a and Fig. 3a). Another verification of the reliability of our semiquantitative method was performed using the measurement of NIST plasma as available standard reference materials to compare the obtained concentration values with literature^22, 23, 24^. Unfortunately, no information about sulfatide concentrations is found in the literature, so that the comparison of obtained concentrations is performed only for SM lipid class in Supplementary Figure 1, showing very good congruence.

### Lipid annotation

The lipid nomenclature followed LIPID MAPS system^52^ and the shorthand notation for lipid structures^53, 54^. In the following text, sulfatide shorthand notification is briefly described on the example of sulfatide SHex2Cer 34:1;O2 for a better understanding. First capital letter S means presence of one sulfate group, the abbreviation Hex means the presence of hexosyl unit without specification of stereochemistry, and the additional number gives the information on their number (in our example, two hexosyl units) on ceramide backbone. Subsequently, the colon-separated numbers 34:1 provides the information on the total number of carbon atoms and double bonds (CN:DB) of N-linked fatty acyl and sphingoid base of ceramide part, and a semicolon separated O2 informs about two hydroxyl oxygens in the ceramide part without any specification of their position. In the case that an additional hydroxyl is present in the ceramide part without any specification of its position O3 is written behind semicolon.

For sterol sulfates, we have been able to verify the presence of the sulfate group according to MS/MS spectra, where a typical neutral loss of 80 was observed, and using a high-resolution mass spectra measurement that allowed us to determine the elemental composition. However, the exact structure elucidation, including the isomerism of sterols, is not possible only by using MALDI-Orbitrap MS measurement. Several sterol isomers with identical elemental composition can contribute to the ion signal. For this reason, we have abbreviated the observed sterols from StS 1 to St S13 and we only suggested the most probable compounds for individual elemental compositions in Supplementary Table 1.

### Statistical data analysis

All calculations and visualizations were done using Microsoft Excel (ver. 2111), R free software environment (ver. 4.1.2), SIMCA statistical software (ver. 13.0.0.0), and the Inkscape open-source vector graphics editor (ver. 1.1). In the first step, the distribution of variables in the dataset was usually observed based on the density plots generated using the ggplot2 library. Then, medians and interquartile ranges (IQR) were calculated for all the variables within groups, using the rstatix library. Fold change was computed *via* the robust Hodges-Lehmann type fold change estimator. To obtain it, we computed all possible pairwise ratios between the feature’s values in two groups, and then the median of those ratios was returned. To compare differences between two independent groups, we used the Mann-Whitney U test (plasma and urine), and its equivalent for paired samples was applied in the case of tissue samples (rstatix package). The comparison of controls and cases split into stages or grades (multiple group analyses) was performed using the Kruskal Wallis test (rstatix library). When the Kruskal-Wallis test indicated differences, the Conover post-hoc test was applied to see which pairs of groups differed significantly (PMCMRplus library). The rank-biserial correlation coefficient effect size was computed for the Mann-Whitney two-sample test using the rcompanion library, and the epsilon squared effect size for the Kruskal-Wallis test (with the use of the ggstatsplot library). For the presence/absence analysis, first, the contingency tables were prepared in which lipids having intensity > 8000 were classified as “1” (present), otherwise as “0” (absent). The Fisher’s exact test was used to analyze contingency tables concerning lipids detected in plasma and urine samples. In the case of tissues, both parts were taken from the same subject, and we considered the following pairs: lipid present (or absent) in nontumor/tumor tissue at once, i.e., (1,1) or (0,0), present only in nontumor tissue (1,0), or only in tumor tissue (0,1). The McNemar’s test was applied in the next step (exact2×2 library) to analyze the contingency tables for tissues. Spearman correlations were calculated to investigate the relationships between age and lipid concentrations and BMI and lipid concentrations (psych library). Volcano plots were prepared with the use of the EnhancedVolcano library. Differences between groups were visualized in the form of box plots for skewed distributions (litteR and ggplot2 libraries). The construction of a box plot was presented and well-explained in Hubert & Vandervieren^55^. All p-values were corrected for the multiple comparisons using the Benjamini-Hochberg assessment of the False Discovery Rate (FDR). In the next step, the data were log-transformed and Pareto-scaled (tidymodels, tidySpectR libraries). The Principal Component Analysis was performed in the SIMCA statistical software. For the circular dendrograms, the Euclidean distances were calculated first, and Ward’s clustering algorithm was used in the next step (phangorn library). The dendrogram was generated applying the ggtree library and the ggplot2 library. The z-scores were calculated for each transformed concentration before plotting the heat map. The logistic regression with ridge penalty was applied to classify plasma samples (glmnet library). All 33 lipid concentrations, presence/absence data for 13 lipids, BMI, and gender were included in the model. The data were split into training (75%) and testing set (25%), log-transformed, and Pareto-scaled. A 5-fold cross-validation approach was applied to prepare the model. Based on the AUC values, the model with a minimum value of lambda regularization parameter was selected for the classification. The receiver operating characteristic curve (ROC) was generated (pROC library) to assess the model performance, and the confusion matrix was computed using the caret library. Confidence intervals were calculated for accuracy, specificity, and sensitivity. The ROC curves were also prepared for individual lipids in all analyzed samples. The AUC from this analysis and fold changes were subsequently used to generate Fig. 6 in the Cytoscape software (the network visualization of alterations in lipid profiles of RCC vs. controls). The color saturation reflects fold change and the size of each circle - the AUC for a lipid.

## Data Availability

All data necessary to support the conclusions are available in the manuscript or supplementary information. Raw data, instructions for data processing and Excel macro script are available in online open access repository Figshare

https://figshare.com/s/d01d43e862b7e03adbfe

## Data availability

All data necessary to support the conclusions are available in the manuscript or supplementary information. Raw data, instructions for data processing and Excel macro script are available in online open-access repository Figshare at https://figshare.com/s/d01d43e862b7e03adbfe

## Acknowledgements

This work was supported by Czech Science Foundation (GAČR) project No. 18-12204S. The authors would like to acknowledge Eva Cífková and Pavel Čáň for the development of Excel macro script for processing of lipidomic data and Ivana Brabcová for the extraction of urine samples.

## Author contributions

RJ, and MH prepared the concept of the study. DW performed the sample preparation. RJ analyzed samples and processed data. Statistical analysis was performed by JI and consulted with KP, DF and AK Coauthors HS, VS and BM obtained and provided plasma samples and clinical information. RJ prepared the first draft of the manuscript. AK, DF, JI contributed to biological discussion. JI and RJ prepared Figures. MH was responsible for funding of this study. All co-authors read, reviewed, edited, and approved the manuscript.

