## Supplementary_Figures_1-13 for "Altered plasma, urine, and tissue profiles of sulfatides and sphingomyelins in patients with renal cell carcinoma"

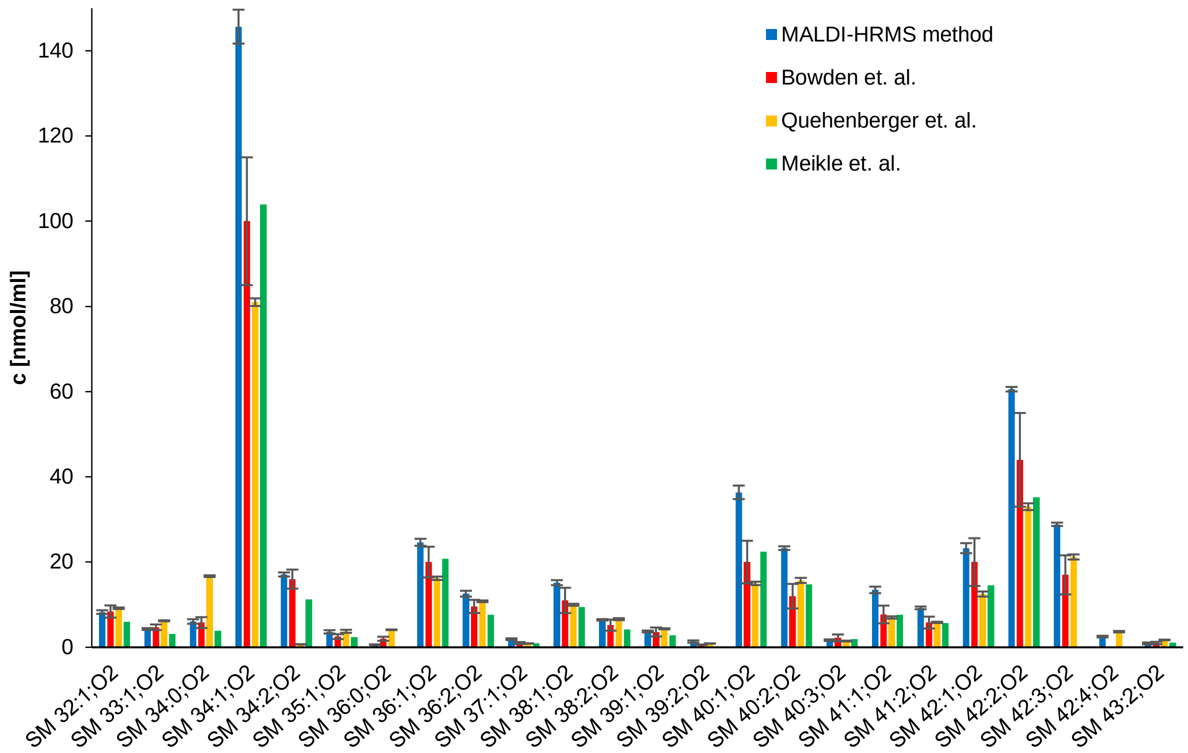


**Fig. 1** Bar graph compares molar concentrations between our MALDI-HRMS method (blue bars, error bars represent standard deviation (SD) and were calculated based on the three independently extracted samples) and other data from the literature[22, 23, 24].

**a**

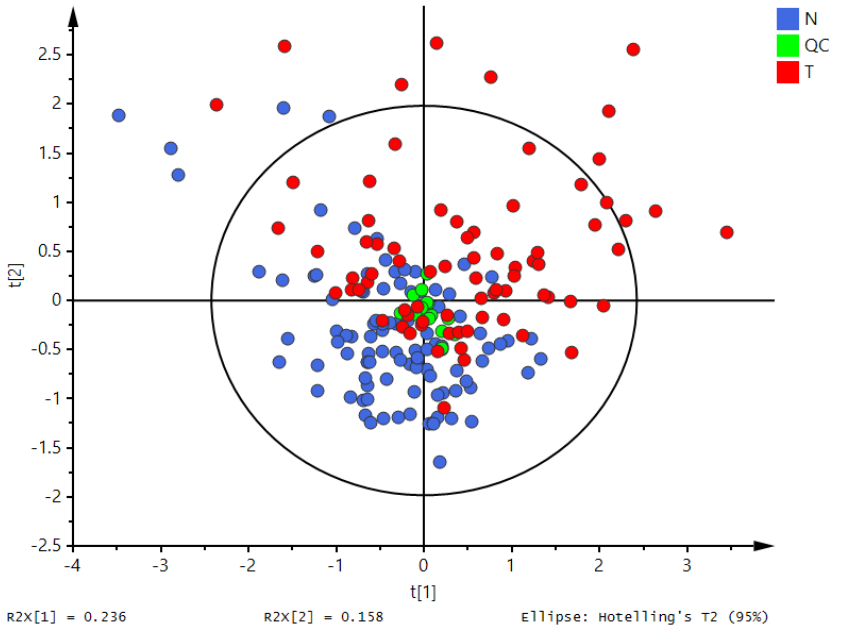


**b**

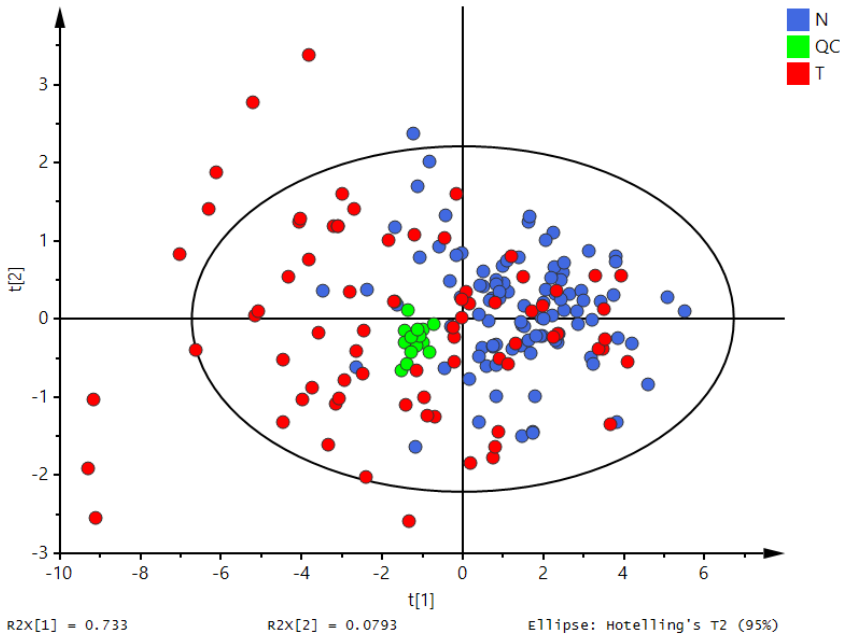


**Fig. 2** PCA-X score plots built based on **a** relative and **b** molar concentrations in plasma samples of 100 controls (N, blue), 75 RCC patients (T, red) and 18 quality control samples (QC, green). In these models, samples obtained from people younger than 49 years and older than 70 years were excluded so the age is balanced between both studied groups. Data were log-transformed and Pareto-scaled before the PCA analysis.

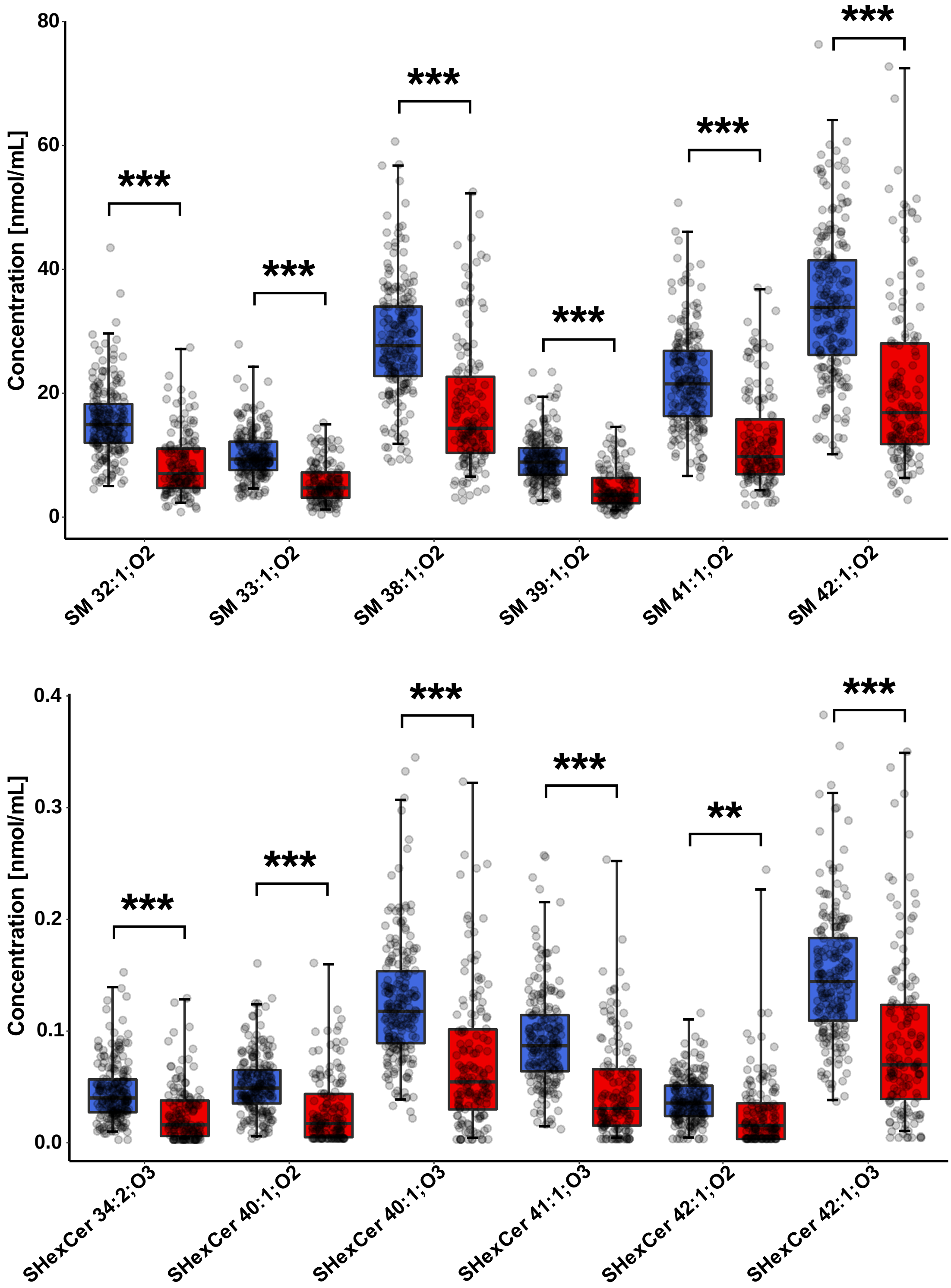


**Fig. 3** Box plots of the molar concentrations for selected statistically significant SHexCer and SM in plasma samples for controls (blue) and RCC patients (red). Box plots were adjusted for skewed distributions according Hubert & Vandervieren[55]. The importance marked above box plots includes fold change, effect size, and FDR from the Mann-Whitney U test: **** - very large, *** - large, ** - medium, * - small significance (more explanation in the Suppl. Data 11).

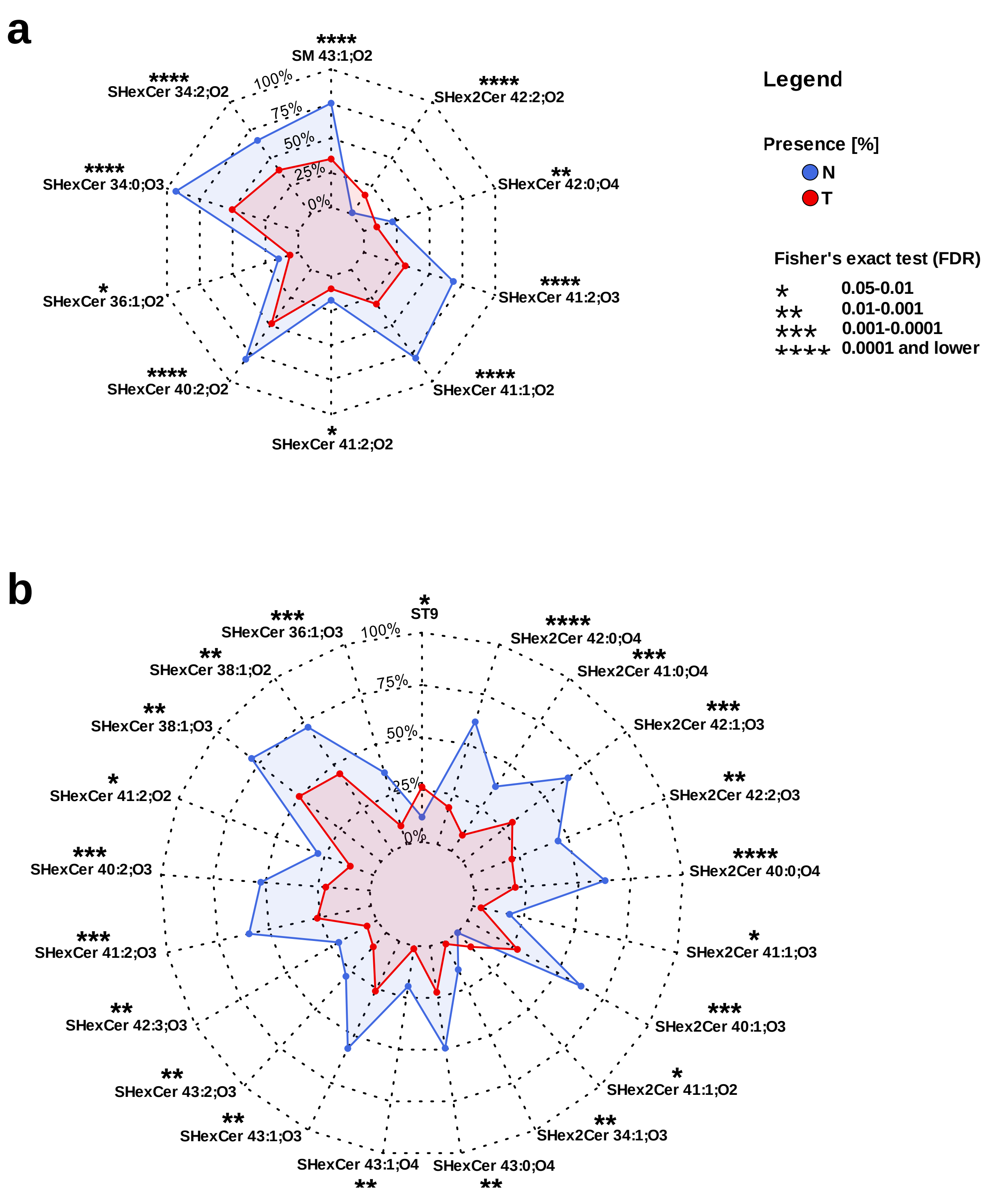


**Fig. 4** Radar charts of SHexCer and SM species analyzed using the present/absent approach for **a** plasma and **b** urine samples of controls (blue) and RCC patients (red). Contingency tables (Supplementary Data 12 and 14) were created for these lipids, and Fisher's exact test was applied for the presence/absence analysis.

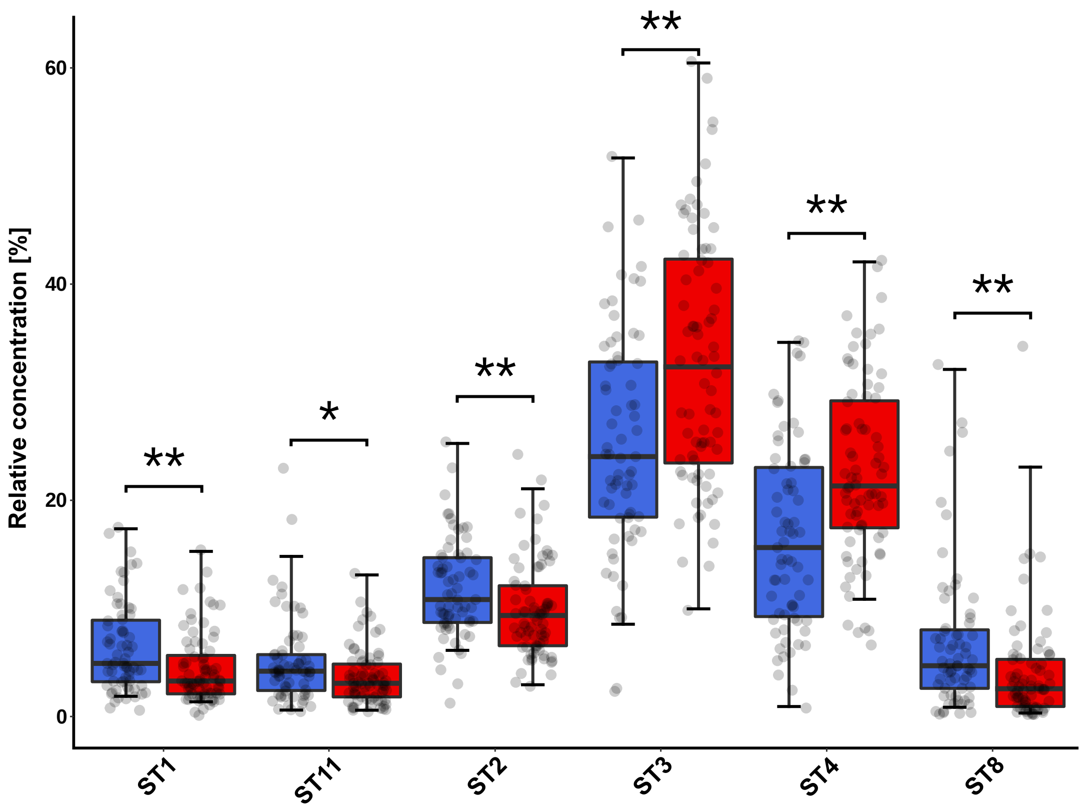


**Fig. 5** Box plots (relative concentrations) of selected sterols measured in urine samples for controls (blue) and patients with RCC (red). Box plots were adjusted for skewed distributions according Hubert & Vandervieren[55]. The importance marked above box plots includes fold change, effect size, and FDR from the Mann-Whitney U test: **** - very large, *** - large, ** - medium, * - small significance (more explanation in the Supplementary Data 13).

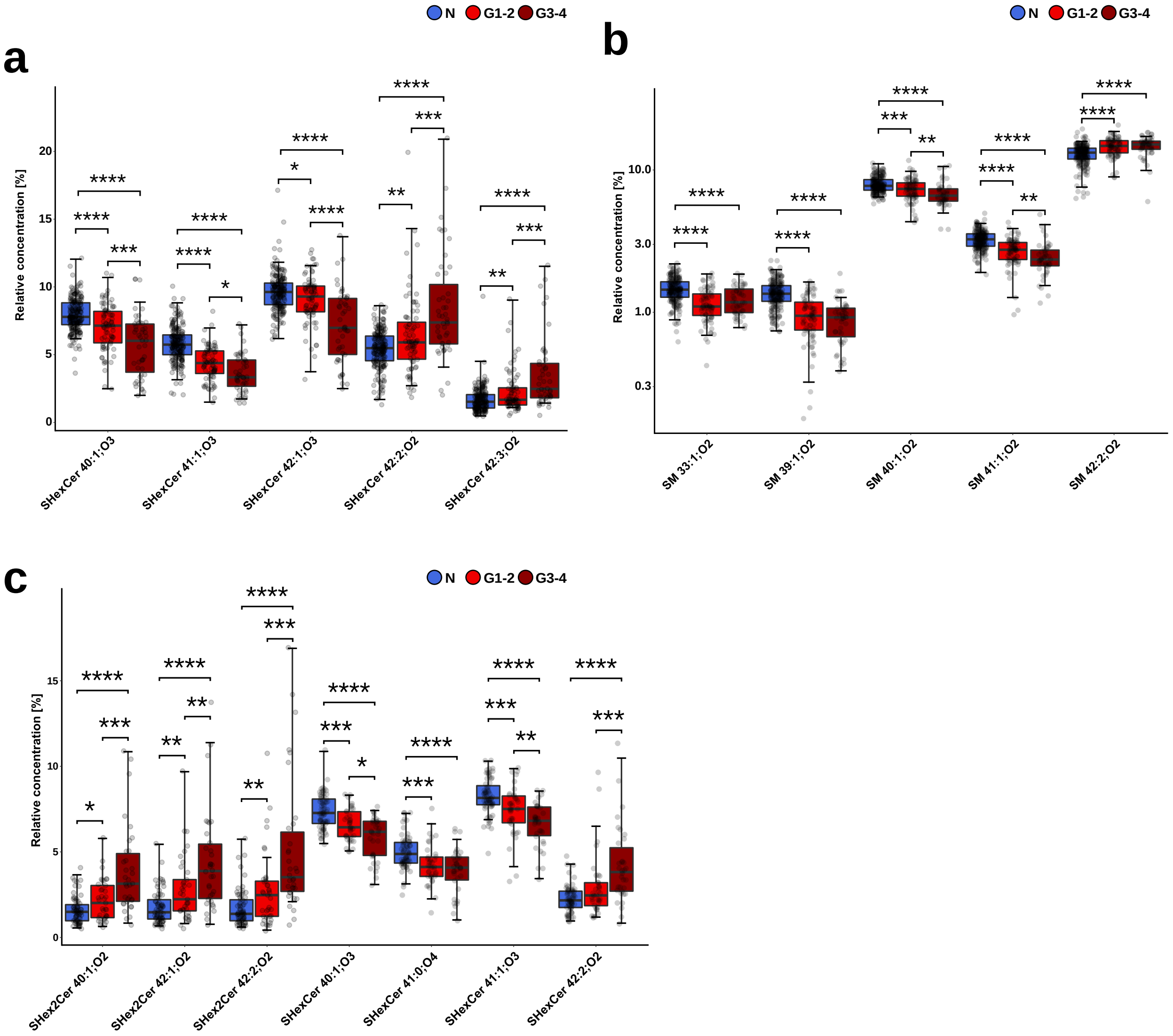


**Fig. 6** Box plots of the most statistically significant lipids for controls, RCC patients with tumor grade G1-2, and RCC patients with tumor grade G3-4: **a** sulfatides in plasma samples, **b** SM in plasma samples, and **c** sulfatides in urine samples. The importance marked above box plots indicates results of the Conover posthoc test (FDR): 0.05-0.01 - *, 0.01-0.001 - **, 0.001-0.0001 - ***, 0.0001 and lower - ****.

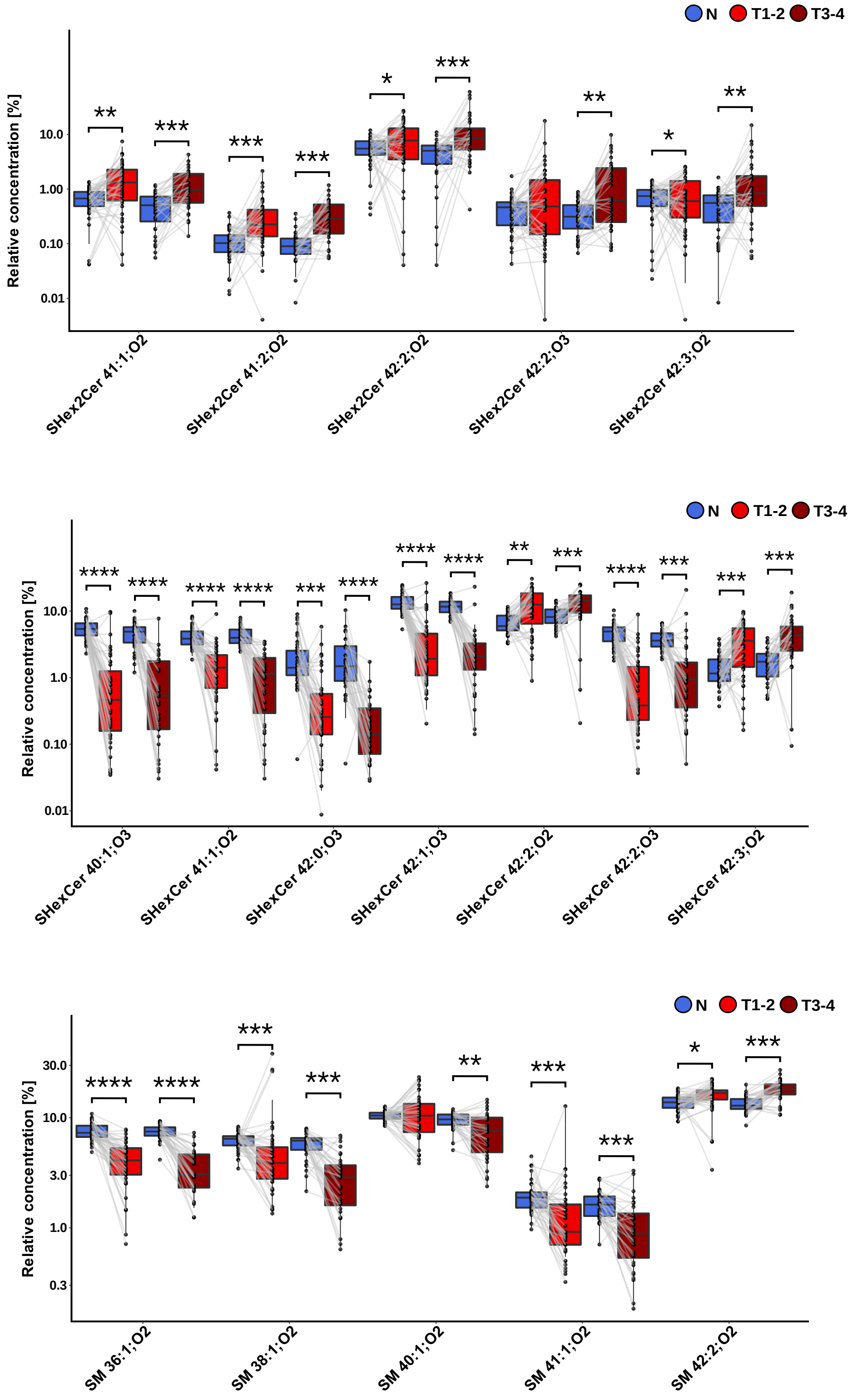


**Fig. 7** Comparison of the most statistically significant lipids in tumor tissue and surrounding nonneoplastic tissue of RCC patients with early tumor stage (T1-T2) and RCC patients with more advanced tumor stage (T3-T4). Box plots were adjusted for skewed distributions according Hubert & Vandervieren[55]. The importance marked above box plots includes fold change, effect size, and FDR from the Wilcoxon signed-rank test: **** - very large, *** - large, ** - medium, * - small significance (more explanation in the Suppl. material 21 & 22 - tissues).

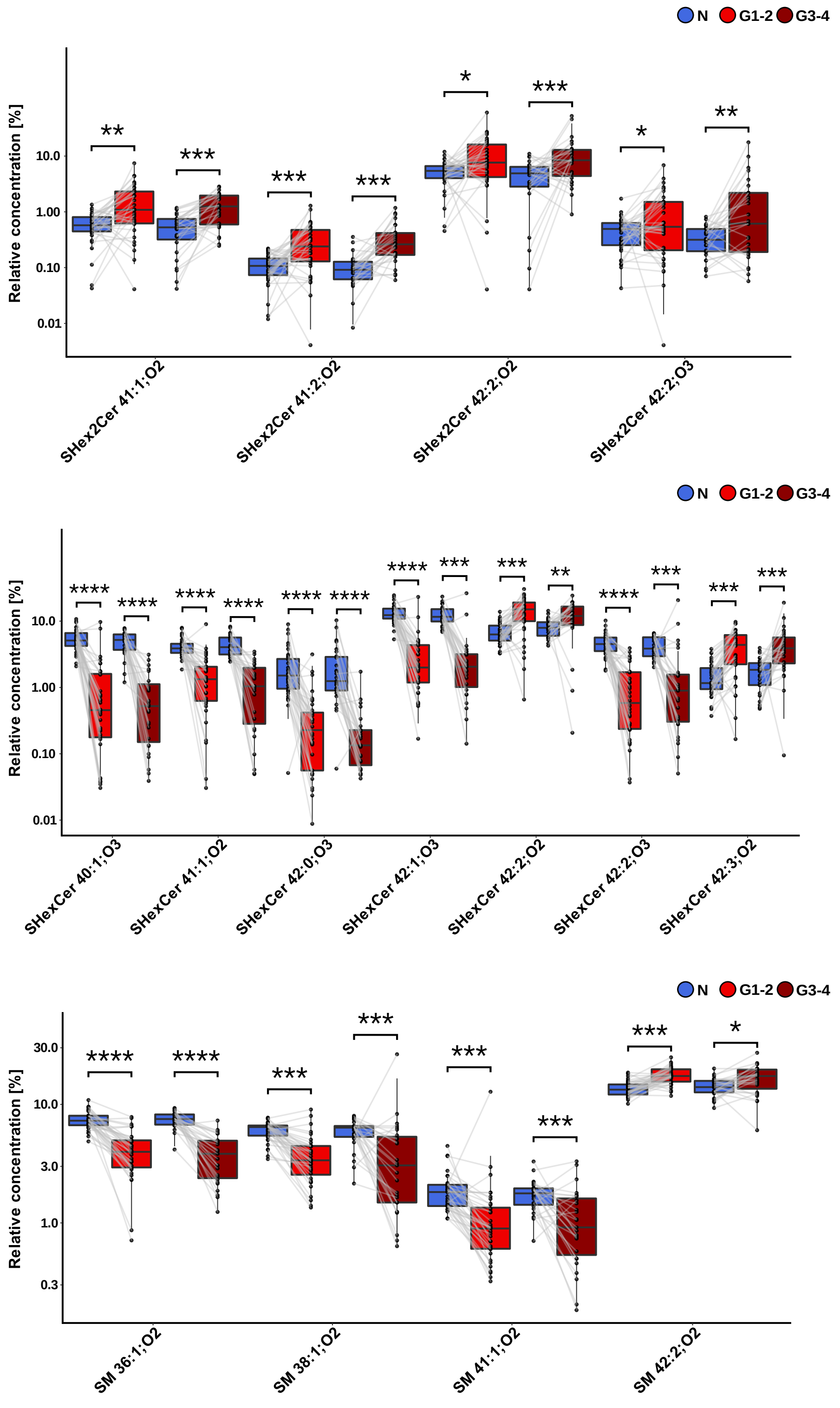


**Fig. 8** Comparison of the most statistically significant lipids in tumor tissue and adjacent nonneoplastic tissue samples of RCC patients with lower tumor grade (G1-G2), and RCC patients with more advanced tumor grades (G3-G4). Box plots were adjusted for skewed distributions according Hubert & Vandervieren[55]. The importance marked above box plots includes fold change, effect size, and FDR from the Wilcoxon signed-rank test: **** - very large, *** - large, ** - medium, * - small significance (more explanation in the Suppl. material 23 & 24 - tissues).

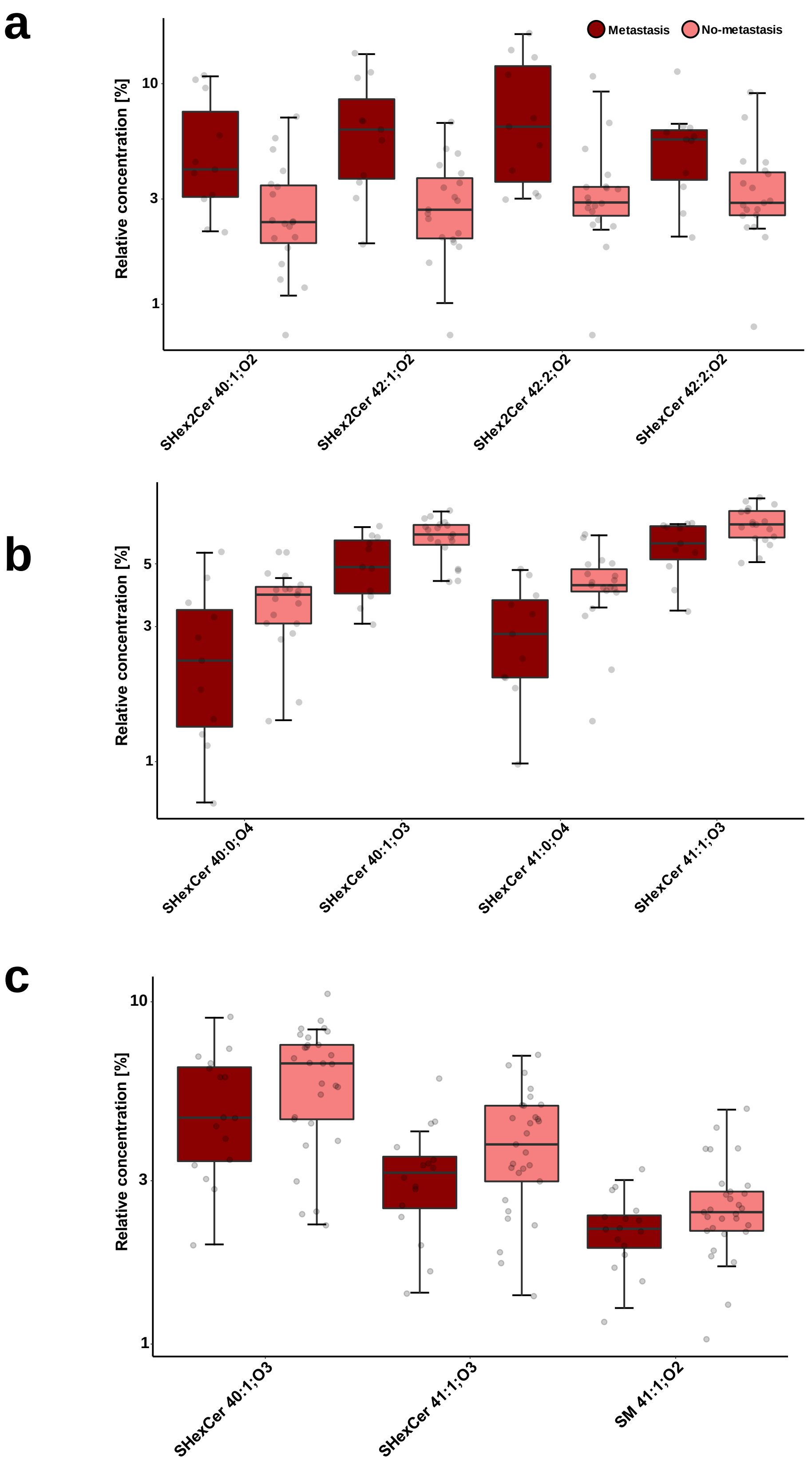


**Fig. 9** Box plots present differences in relative concentrations of selected lipids between RCC patients with T3 tumor stage with (dark red) or without metastases (pink), measured in urine (**a**, **b**) and plasma (**c**). Box plots were adjusted for skewed distributions according Hubert & Vandervieren[55].

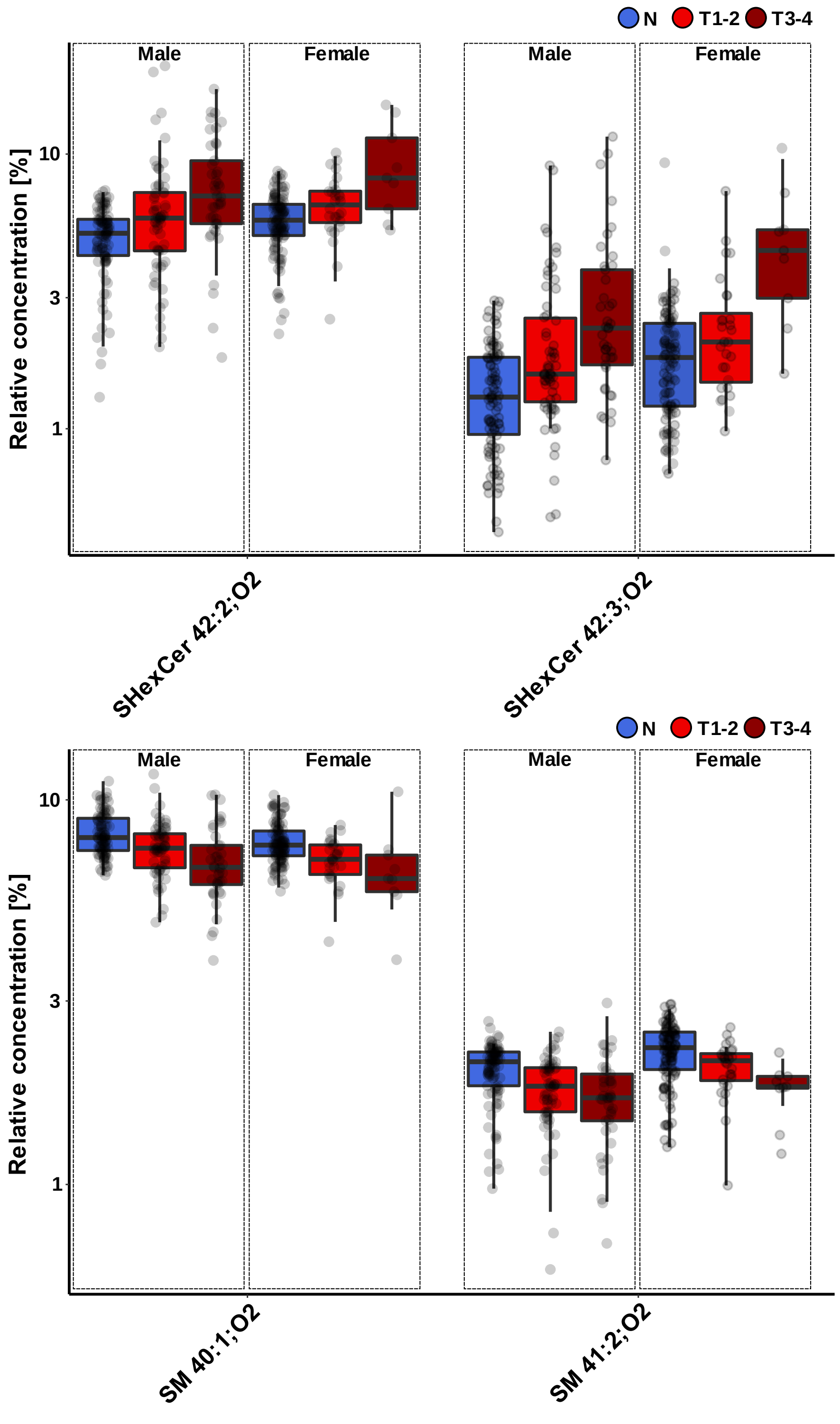


**Fig. 10** Box plots present subtle differences in the relative concentration of selected lipids between genders in studied plasma samples (N = controls, T1-2 = patients with early tumor stage, T3-4 = patients with more advanced tumor stage). Box plots were adjusted for skewed distributions according Hubert & Vandervieren[55].

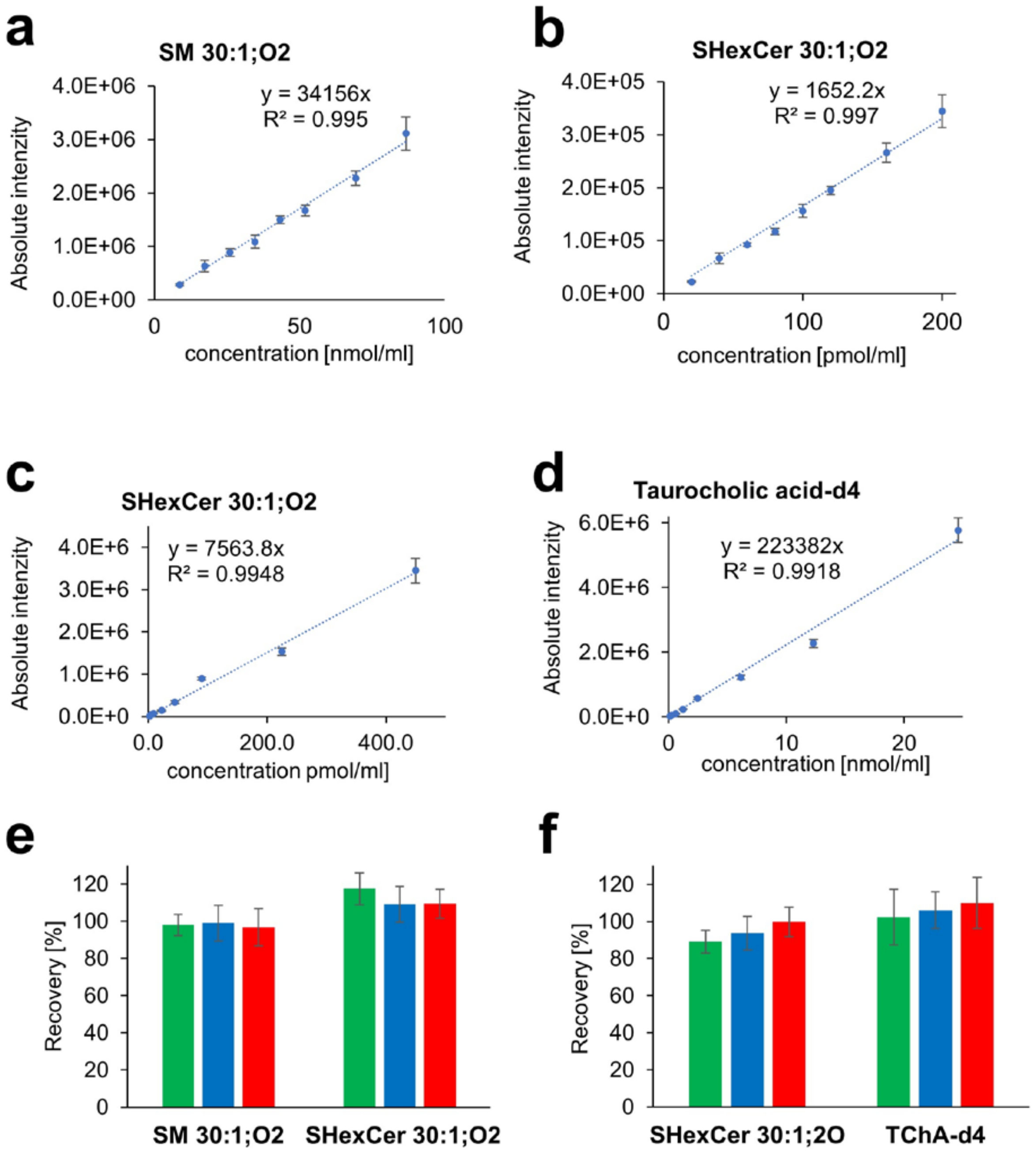


**Fig. 11** Calibration curves present linear concentration ranges for applied internal standards in (a, b) plasma and (c, d) urine samples (error bars represent SD from five consecutive measurements). Extraction recovery of internal standards used for the extraction of (e) plasma and (f) urine samples (error bars represent the coefficient of variation of 6 independent extractions in the case of plasma and 3 independent extractions in the case of urine).

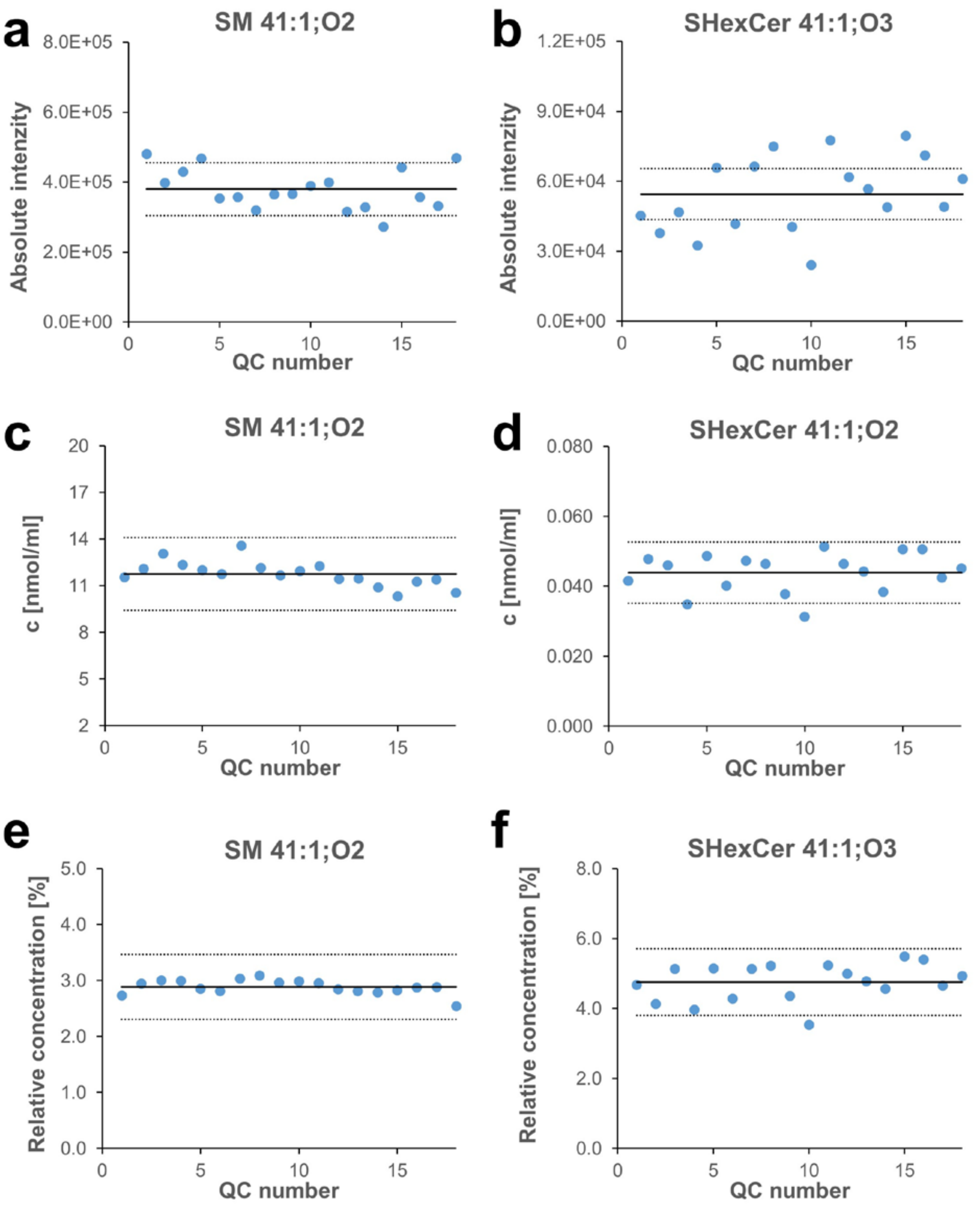


**Fig. 12** Quality control of lipidomic analysis based on the response of 18 QC samples during the sequence measurement of plasma samples in the example of endogenously present lipids SM 41:1;O2 and SHexCer 41:1;O3: **a, b** absolute intensities; **c, d** calculated molar concentrations; **e, f** relative concentrations. The middle full lines represent the average of individual values (intensities, molar, or relative concentrations), and two dotted lines are twenty percentage deviations from this average value.

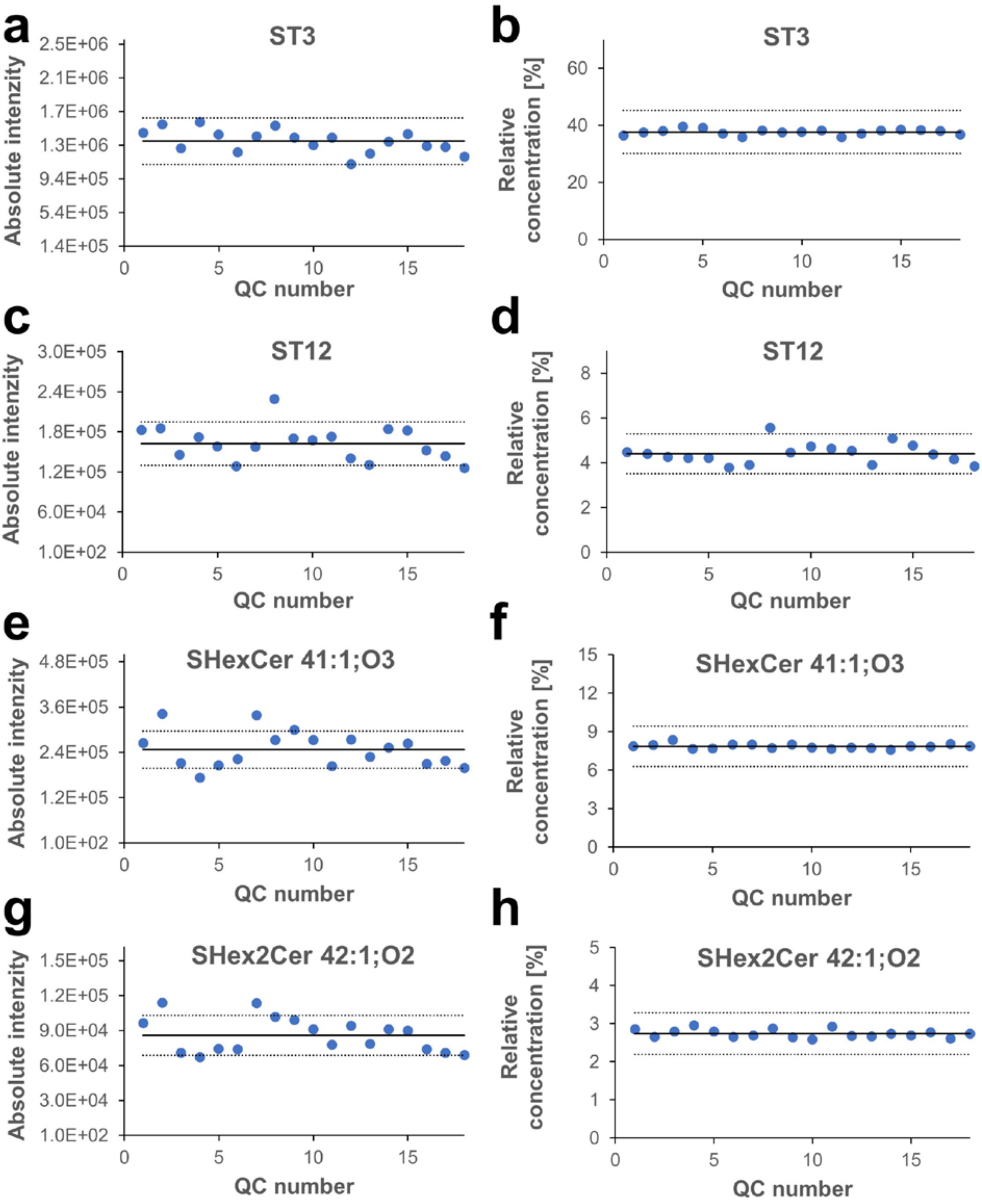


**Fig. 13** Quality control of lipidomic analysis based on the response signals of QC samples during the sequence measurement of urine samples in the example of endogenously present lipids ST3, ST12, SHexCer 41:1;O3 and SHex2Cer 42:1;O2: **a, c, e, g** absolute intensities; **b, d, f, h** relative concentrations. The middle full lines represent the average of individual values (intensities or relative concentrations), and two dotted lines are twenty percentage deviations from this average value.
