## Supplementary_Tables_1-12 for "Altered plasma, urine, and tissue profiles of sulfatides and sphingomyelins in patients with renal cell carcinoma"

**Table 1** Abbreviation of the sterols observed in urine samples with theoretical m/z value, elemental composition, and suggestion of the most probable corresponding compounds for individual elemental compositions.

| Abbreviation | Theoretical m/z of $[M-H]^-$ | Elemental composition | Suggested compound |
|---|---|---|---|
| StS 1 | 411.1847 | $C_{21}H_{32}O_6S$ | tetrahydrocorticosterone sulfate -$H_2O$ |
| StS 2 | 429.1953 | $C_{21}H_{34}O_7S$ | tetrahydrocorticosterone sulfate |
| StS 3 | 441.1589 | $C_{21}H_{30}O8S$ | cortisol sulfate |
| StS 4 | 445.1902 | $C_{21}H_{34}O_8S$ | tetrahydrocortisol sulfate |
| StS 5 | 455.2473 | $C_{24}H_{40}O_6S$ | lithocholic acid sulfate |
| StS 6 | 465.304 | $C_{27}H_{46}O_4S$ | cholesterol sulfate |
| StS 7 | 471.2422 | $C_{24}H_{40}O_7S$ | ursodeoxycholic acid sulfate |
| StS 8 | 482.2946 | $C_{26}H_{45}NO_5S$ | taurolithocholic acid |
| StS 9 | 487.2371 | $C_{24}H_{40}O_8S$ | sulfocholic acid |
| StS 10 | 498.2895 | $C_{26}H_{45}NO_6S$ | taurodeoxycholic acid |
| StS 11 | 512.2688 | $C_{26}H_{43}NO_7S$ | sulfoglycolithocholic acid |
| StS 12 | 514.2844 | $C_{26}H_{45}NO_7S$ | taurocholic acid |
| StS 13 | 528.2637 | $C_{26}H_{43}NO_8S$ | glycochenodeoxycholic acid sulfate |

**Table 2** Spearman correlation heatmap showing values of positive (red) and negative (blue) correlations between individual lipids studied and age in plasma samples from controls (N), patients with tumor stage T1-2, and patients with tumor stage T3-4, separately shown for males and females. For values in bold FDR < 0.05, and stronger saturation reflects higher association.

| | | **Females** | | | **Males** | | |
|---|---|---|---|---|---|---|---|
| | | **N** | **T1-2** | **T3-4** | **N** | **T1-2** | **T3-4** |
| Age | SM 32:1;O2 | 0.03 | -0.05 | 0.08 | -0.08 | 0.11 | **0.45** |
| Age | SM 33:1;O2 | -0.09 | 0.01 | 0.25 | -0.06 | 0.19 | 0.32 |
| Age | SM 34:2;O2 | **0.43** | 0.17 | 0.32 | **0.29** | 0.09 | 0.21 |
| Age | SM 34:1;O2 | -0.21 | 0.14 | 0.33 | 0.00 | **0.39** | 0.30 |
| Age | SM 34:0;O2 | **-0.22** | -0.08 | -0.13 | 0.00 | 0.24 | 0.25 |
| Age | SM 35:1;O2 | -0.05 | -0.16 | -0.18 | -0.17 | 0.27 | 0.35 |
| Age | SM 36:2;O2 | **0.40** | 0.10 | 0.50 | 0.19 | 0.00 | 0.09 |
| Age | SM 36:1;O2 | 0.08 | -0.17 | 0.30 | 0.01 | 0.06 | -0.06 |
| Age | SM 36:0;O2 | 0.20 | -0.06 | -0.10 | -0.04 | -0.06 | -0.04 |
| Age | SM 37:1;O2 | 0.04 | -0.19 | -0.13 | -0.10 | 0.00 | 0.14 |
| Age | SM 38:2;O2 | **0.29** | 0.10 | 0.18 | **0.27** | -0.04 | -0.02 |
| Age | SM 38:1;O2 | -0.14 | 0.07 | -0.40 | -0.05 | -0.22 | -0.09 |
| Age | SM 39:1;O2 | -0.02 | 0.11 | -0.33 | -0.11 | -0.16 | 0.09 |
| Age | SM 40:2;O2 | **0.27** | -0.04 | -0.03 | 0.06 | -0.27 | 0.06 |
| Age | SM 40:1;O2 | **-0.25** | -0.02 | -0.63 | -0.12 | **-0.38** | -0.24 |
| Age | SM 41:2;O2 | 0.18 | 0.11 | -0.17 | 0.04 | -0.11 | 0.31 |
| Age | SM 41:1;O2 | -0.15 | 0.07 | -0.37 | -0.20 | -0.27 | -0.02 |
| Age | SM 42:3;O2 | **0.22** | 0.06 | 0.17 | 0.12 | -0.01 | -0.07 |
| Age | SM 42:2;O2 | 0.04 | 0.00 | -0.40 | 0.04 | -0.04 | -0.36 |
| Age | SM 42:1;O2 | **-0.24** | -0.07 | -0.50 | **-0.24** | **-0.36** | -0.25 |
| Age | SM 43:2;O2 | **0.23** | -0.02 | -0.25 | 0.00 | 0.11 | 0.06 |
| Age | SHexCer 34:1;O2 | **0.28** | -0.04 | 0.80 | 0.06 | 0.19 | **0.42** |
| Age | SHexCer 34:1;O3 | **-0.30** | 0.13 | 0.17 | 0.01 | 0.11 | 0.27 |
| Age | SHexCer 40:1;O2 | **0.26** | -0.01 | -0.72 | -0.02 | -0.05 | -0.02 |
| Age | SHexCer 40:2;O3 | 0.12 | 0.05 | -0.15 | 0.04 | -0.07 | -0.09 |
| Age | SHexCer 40:1;O3 | -0.07 | -0.22 | -0.23 | -0.14 | -0.32 | -0.07 |
| Age | SHexCer 42:3;O2 | -0.13 | 0.14 | -0.02 | 0.02 | 0.09 | **-0.43** |
| Age | SHexCer 42:2;O2 | -0.07 | 0.27 | -0.52 | -0.08 | 0.17 | -0.28 |
| Age | SHexCer 42:1;O2 | 0.11 | 0.02 | -0.55 | 0.02 | 0.08 | -0.26 |
| Age | SHexCer 41:1;O3 | 0.02 | -0.37 | -0.40 | -0.17 | -0.26 | 0.06 |
| Age | SHexCer 42:3;O3 | 0.20 | 0.10 | -0.12 | 0.03 | 0.00 | -0.14 |
| Age | SHexCer 42:2;O3 | 0.02 | 0.02 | -0.03 | 0.05 | -0.23 | -0.26 |
| Age | SHexCer 42:1;O3 | -0.08 | 0.00 | -0.27 | 0.01 | -0.41 | -0.04 |

**Table 3** Spearman correlation heatmap showing values of positive (red) and negative (blue) correlations between individual lipids studied and BMI in plasma samples from controls (N), patients with tumor stage T1-2, and patients with tumor stage T3-4, shown separately for males and females. For values in bold FDR < 0.05, and stronger saturation reflects higher association.

| | | Females | | | Males | | |
|---|---|---|---|---|---|---|---|
| | | N | T1-2 | T3-4 | N | T1-2 | T3-4 |
| BMI | SM 32:1;O2 | 0.08 | -0.02 | 0.69 | 0.03 | -0.03 | 0.02 |
| BMI | SM 33:1;O2 | 0.00 | -0.37 | 0.50 | -0.23 | 0.01 | 0.02 |
| BMI | SM 34:2;O2 | **0.28** | 0.52 | 0.64 | -0.06 | 0.07 | 0.20 |
| BMI | SM 34:1;O2 | **-0.25** | -0.39 | -0.40 | **-0.32** | -0.31 | -0.32 |
| BMI | SM 34:0;O2 | -0.18 | -0.30 | -0.52 | 0.07 | -0.05 | 0.09 |
| BMI | SM 35:1;O2 | 0.11 | -0.26 | 0.17 | 0.05 | -0.12 | 0.18 |
| BMI | SM 36:2;O2 | **0.34** | 0.52 | 0.48 | 0.19 | 0.29 | 0.23 |
| BMI | SM 36:1;O2 | 0.22 | 0.31 | -0.21 | **0.47** | **0.39** | 0.07 |
| BMI | SM 36:0;O2 | **0.44** | 0.39 | -0.12 | **0.44** | 0.32 | 0.05 |
| BMI | SM 37:1;O2 | **0.37** | -0.11 | 0.33 | **0.36** | 0.11 | 0.03 |
| BMI | SM 38:2;O2 | 0.22 | 0.32 | 0.43 | 0.02 | 0.20 | 0.29 |
| BMI | SM 38:1;O2 | 0.10 | -0.10 | 0.52 | **0.36** | **0.43** | 0.24 |
| BMI | SM 39:1;O2 | 0.15 | -0.25 | 0.67 | 0.13 | 0.20 | 0.31 |
| BMI | SM 40:2;O2 | 0.15 | 0.39 | 0.48 | 0.03 | 0.06 | 0.10 |
| BMI | SM 40:1;O2 | 0.11 | 0.13 | 0.14 | **0.36** | **0.33** | 0.35 |
| BMI | SM 41:2;O2 | 0.04 | -0.21 | 0.76 | -0.12 | -0.15 | 0.13 |
| BMI | SM 41:1;O2 | 0.08 | -0.16 | 0.64 | 0.17 | 0.22 | **0.44** |
| BMI | SM 42:3;O2 | 0.05 | 0.33 | 0.19 | **-0.24** | -0.30 | 0.00 |
| BMI | SM 42:2;O2 | -0.11 | -0.08 | -0.60 | -0.15 | -0.21 | -0.35 |
| BMI | SM 42:1;O2 | 0.03 | 0.02 | 0.07 | 0.20 | 0.10 | 0.30 |
| BMI | SM 43:2;O2 | 0.12 | -0.40 | -0.14 | -0.01 | -0.13 | -0.21 |
| BMI | SHexCer 34:1;O2 | 0.11 | 0.08 | 0.31 | 0.11 | 0.22 | 0.01 |
| BMI | SHexCer 34:1;O3 | -0.07 | 0.18 | -0.10 | -0.03 | 0.17 | 0.07 |
| BMI | SHexCer 40:1;O2 | 0.19 | -0.01 | 0.17 | 0.06 | -0.11 | 0.39 |
| BMI | SHexCer 40:2;O3 | 0.13 | 0.06 | -0.10 | -0.22 | -0.10 | 0.05 |
| BMI | SHexCer 40:1;O3 | **0.27** | 0.16 | 0.74 | 0.19 | 0.21 | 0.27 |
| BMI | SHexCer 42:3;O2 | **-0.40** | -0.01 | -0.60 | **-0.46** | **-0.37** | -0.15 |
| BMI | SHexCer 42:2;O2 | **-0.43** | -0.30 | -0.67 | **-0.39** | **-0.54** | -0.33 |
| BMI | SHexCer 42:1;O2 | **-0.25** | -0.25 | -0.64 | -0.14 | -0.31 | 0.14 |
| BMI | SHexCer 41:1;O3 | 0.12 | -0.46 | 0.48 | -0.05 | -0.18 | 0.15 |
| BMI | SHexCer 42:3;O3 | 0.07 | -0.13 | -0.57 | **-0.24** | -0.19 | -0.05 |
| BMI | SHexCer 42:2;O3 | 0.11 | 0.27 | 0.69 | -0.03 | 0.07 | -0.12 |
| BMI | SHexCer 42:1;O3 | 0.13 | -0.18 | 0.57 | **0.31** | 0.28 | 0.28 |

**Table 4** List of concentrations of individual internal standards used for the analytical validation experiments in plasma and urine samples for low (LL), medium (ML), and high (HL) concentration levels.

| | **c [nmol/ml of plasma]** | | |
|---|---|---|---|
| | **LL** | **ML** | **HL** |
| SM 18:1;O2/12:0 | 21.65 | 43.3 | 64.95 |
| SHexCer 18:1;O2/12:0 | 0.05 | 0.1 | 0.15 |

| | **c [nmol/ml of urine]** | | |
|---|---|---|---|
| | **LL** | **ML** | **HL** |
| Taurocholic acid D4 | 0.183 | 0.55 | 1.65 |
| SHexCer 18:1;O2/12:0 | 0.013 | 0.04 | 0.12 |

**Table 5** Median of absolute intensities and individual coefficients of variation (CV) for each IS - calculated from 5 repetitions (consecutive measurements) within individual samples of the plasma validation experiment. Data have been used to calculate recovery rates (x = IS added before the extraction and y = IS added after extraction) and precisions (x). The individual CV values represent repeatability.

| IS addition | Day | Level | SM 30:1;O2 | | SHexCer 30:1;O2 | |
|---|---|---|---|---|---|---|
| | | | Median of Intensity | CV [%] | Median of Intensity | CV [%] |
| Low level (IS added before extraction) | 1st day | Lx1 | 8.25E+05 | 7 | 1.00E+05 | 19 |
| | | Lx2 | 7.62E+05 | 5 | 8.94E+04 | 12 |
| | | Lx3 | 7.57E+05 | 8 | 8.15E+04 | 14 |
| | 2nd day | Lx4 | 7.76E+05 | 7 | 1.05E+05 | 4 |
| | | Lx5 | 8.20E+05 | 7 | 9.94E+04 | 6 |
| | | Lx6 | 8.29E+05 | 14 | 1.04E+05 | 4 |
| Low level (IS added after extraction) | 1st day | Ly1 | 7.83E+05 | 10 | 7.72E+04 | 11 |
| | | Ly2 | 7.45E+05 | 14 | 7.36E+04 | 10 |
| | | Ly3 | 7.65E+05 | 12 | 6.74E+04 | 12 |
| | 2nd day | Ly4 | 8.59E+05 | 8 | 1.03E+05 | 5 |
| | | Ly5 | 9.05E+05 | 4 | 7.98E+04 | 2 |
| | | Ly6 | 8.27E+05 | 7 | 9.78E+04 | 9 |
| Medium level (IS added before extraction) | 1st day | Mx1 | 1.09E+06 | 5 | 1.12E+05 | 16 |
| | | Mx2 | 9.78E+05 | 14 | 1.15E+05 | 10 |
| | | Mx3 | 1.05E+06 | 8 | 1.14E+05 | 2 |
| | 2nd day | Mx4 | 9.10E+05 | 5 | 1.34E+05 | 8 |
| | | Mx5 | 1.13E+06 | 9 | 1.36E+05 | 10 |
| | | Mx6 | 1.03E+06 | 4 | 1.38E+05 | 8 |
| Medium level (IS added after extraction) | 1st day | My1 | 1.09E+06 | 10 | 9.04E+04 | 5 |
| | | My2 | 1.04E+06 | 10 | 9.92E+04 | 11 |
| | | My3 | 9.07E+05 | 7 | 9.96E+04 | 10 |
| | 2nd day | My4 | 1.07E+06 | 7 | 1.40E+05 | 7 |
| | | My5 | 1.08E+06 | 8 | 1.42E+05 | 9 |
| | | My6 | 1.08E+06 | 5 | 1.27E+05 | 9 |
| High level (IS added before extraction) | 1st day | Hx1 | 1.67E+06 | 6 | 2.19E+05 | 5 |
| | | Hx2 | 2.06E+06 | 14 | 2.32E+05 | 9 |
| | | Hx3 | 1.76E+06 | 17 | 2.28E+05 | 16 |
| | 2nd day | Hx4 | 1.71E+06 | 11 | 2.44E+05 | 5 |
| | | Hx5 | 1.91E+06 | 19 | 2.42E+05 | 9 |
| | | Hx6 | 1.77E+06 | 6 | 2.36E+05 | 9 |
| High level (IS added after extraction) | 1st day | Hy1 | 1.92E+06 | 18 | 1.97E+05 | 17 |
| | | Hy2 | 1.78E+06 | 12 | 1.99E+05 | 7 |
| | | Hy3 | 1.90E+06 | 12 | 2.00E+05 | 20 |
| | 2nd day | Hy4 | 1.90E+06 | 7 | 2.05E+05 | 5 |
| | | Hy5 | 1.86E+06 | 10 | 2.36E+05 | 14 |
| | | Hy6 | 1.92E+06 | 10 | 2.52E+05 | 6 |

**Table 6** Median of absolute intensities and individual coefficients of variation (CV) for each IS - calculated from 5 repetitions (consecutive measurements) within individual samples of plasma validation experiments. Data obtained for samples "before freeze/thaw" were used for matrix effect evaluation at three concentration levels. The freeze/thaw stability was evaluated based on the three freeze/thaw cycles for the medium concentration level. The individual CV values represent repeatability.

| Concentration level | Cyclus | Sample No. | SM 30:1;O2 | | SHexCer 30:1;O2 | |
|---|---|---|---|---|---|---|
| | | | Median of Intensity | CV [%] | Median of Intensity | CV [%] |
| Low level | before freeze/thaw | L_31 | 9.08E+05 | 3 | 9.42E+04 | 9 |
| | | L_43 | 8.57E+05 | 8 | 8.69E+04 | 11 |
| | | L_96 | 8.00E+05 | 7 | 8.26E+04 | 9 |
| | | L_97 | 9.50E+05 | 12 | 8.15E+04 | 8 |
| | | L_101 | 1.17E+06 | 18 | 8.78E+04 | 5 |
| | | L_107 | 9.13E+05 | 6 | 7.09E+04 | 13 |
| High level | before freeze/thaw | H_31 | 1.81E+06 | 9 | 2.24E+05 | 2 |
| | | H_43 | 1.66E+06 | 6 | 1.68E+05 | 11 |
| | | H_96 | 1.51E+06 | 8 | 1.67E+05 | 4 |
| | | H_97 | 1.94E+06 | 6 | 1.82E+05 | 17 |
| | | H_101 | 2.49E+06 | 9 | 2.28E+05 | 8 |
| | | H_107 | 2.03E+06 | 5 | 1.99E+05 | 6 |
| Medium level | before freeze/thaw | M_31 | 1.35E+06 | 4 | 1.34E+05 | 7 |
| | | M_43 | 1.01E+06 | 5 | 1.06E+05 | 9 |
| | | M_96 | 1.04E+06 | 11 | 1.05E+05 | 13 |
| | | M_97 | 1.02E+06 | 9 | 1.08E+05 | 16 |
| | | M_101 | 1.40E+06 | 11 | 1.11E+05 | 3 |
| | | M_107 | 1.14E+06 | 5 | 8.73E+04 | 5 |
| | 1st freeze/thaw cyclus | M_31 | 1.03E+06 | 8 | 9.81E+04 | 3 |
| | | M_43 | 8.94E+05 | 8 | 9.35E+04 | 17 |
| | | M_96 | 8.52E+05 | 9 | 8.43E+04 | 3 |
| | | M_97 | 8.49E+05 | 12 | 8.38E+04 | 14 |
| | | M_101 | 1.14E+06 | 10 | 8.62E+04 | 6 |
| | | M_107 | 1.29E+06 | 9 | 9.62E+04 | 21 |
| | 2nd freeze/thaw cyclus | M_31 | 8.81E+05 | 18 | 8.92E+04 | 14 |
| | | M_43 | 8.96E+05 | 10 | 7.19E+04 | 22 |
| | | M_96 | 1.10E+06 | 6 | 1.09E+05 | 14 |
| | | M_97 | 9.27E+05 | 7 | 8.61E+04 | 16 |
| | | M_101 | 1.09E+06 | 9 | 7.96E+04 | 23 |
| | | M_107 | 1.17E+06 | 9 | 1.04E+05 | 23 |
| | 3rd freeze/thaw cyclus | M_31 | 8.83E+05 | 5 | 8.98E+04 | 10 |
| | | M_43 | 1.33E+06 | 13 | 1.42E+05 | 8 |
| | | M_96 | 8.63E+05 | 4 | 7.86E+04 | 11 |
| | | M_97 | 7.78E+05 | 11 | 6.87E+04 | 20 |
| | | M_101 | 1.03E+06 | 3 | 5.67E+04 | 18 |
| | | M_107 | 8.44E+05 | 7 | 6.88E+04 | 10 |

**Table 7** Median of absolute intensities and individual coefficients of variation (CV) for IS used in the case of urine samples - calculated from 5 repetitions within the individual sample type of urine validation experiment. Data have been used for the calculation of recovery rates (x = IS added before the extraction) and y = IS added after extraction), precisions (x), and freeze/thaw stability. The individual CV values represent repeatability.

| **Concentration level (IS addition)** | **Day** | **Level** | **SHexCer 30:1;O2** | | **Taurocholic acid-d4** | |
|---|---|---|---|---|---|---|
| | | | **Median of Intensity** | **CV [%]** | **Median of Intensity** | **CV [%]** |
| Low level (IS added before extraction) | 1st day | Lx1 | 2.72E+04 | 16 | 1.84E+04 | 4 |
| | | Lx2 | 2.43E+04 | 24 | 1.64E+04 | 7 |
| | | Lx3 | 3.42E+04 | 12 | 1.28E+04 | 3 |
| | 2nd day | Lx4 | 2.67E+04 | 21 | 1.81E+04 | 22 |
| | | Lx5 | 2.57E+04 | 14 | 1.98E+04 | 21 |
| | | Lx6 | 1.55E+04 | 23 | 9.80E+03 | 34 |
| Low level (IS added after extraction) | 1st day | Ly1 | 3.20E+04 | 10 | 1.55E+04 | 7 |
| | | Ly2 | 2.88E+04 | 13 | 1.74E+04 | 8 |
| | | Ly3 | 3.50E+04 | 14 | 1.49E+04 | 11 |
| Medium level (IS added before extraction) | 1st day | Mx1 | 1.06E+05 | 9 | 6.60E+04 | 3 |
| | | Mx2 | 7.54E+04 | 15 | 5.38E+04 | 16 |
| | | Mx3 | 9.22E+04 | 12 | 7.86E+04 | 3 |
| Medium level (IS added after extraction) | 1st day | My1 | 1.01E+05 | 19 | 7.64E+04 | 11 |
| | | My2 | 9.10E+04 | 9 | 5.51E+04 | 12 |
| | | My3 | 9.89E+04 | 13 | 6.55E+04 | 8 |
| High level (IS added before extraction) | 1st day | Hx1 | 4.83E+05 | 19 | 3.15E+05 | 2 |
| | | Hx2 | 5.52E+05 | 5 | 3.03E+05 | 4 |
| | | Hx3 | 4.73E+05 | 15 | 2.98E+05 | 11 |
| | 2nd day | Hx4 | 5.05E+05 | 16 | 2.71E+05 | 13 |
| | | Hx5 | 5.44E+05 | 13 | 2.46E+05 | 10 |
| | | Hx6 | 5.11E+05 | 11 | 3.28E+05 | 14 |
| High level (IS added after extraction) | 1st day | Hy1 | 3.87E+05 | 12 | 2.40E+05 | 13 |
| | | Hy2 | 3.37E+05 | 12 | 1.97E+05 | 5 |
| | | Hy3 | 3.72E+05 | 21 | 2.78E+05 | 5 |
| freeze/thaw cyclus | 3rd day | Lx4 | 3.25E+04 | 20 | 1.30E+04 | 16 |
| | | Lx5 | 2.74E+04 | 24 | 1.29E+04 | 12 |
| | | Lx6 | 1.75E+04 | 33 | 9.91E+03 | 30 |
| freeze/thaw cyclus | 3rd day | Hx4 | 3.42E+05 | 15 | 1.82E+05 | 10 |
| | | Hx5 | 3.48E+05 | 14 | 2.20E+05 | 5 |
| | | Hx6 | 2.80E+05 | 11 | 2.55E+05 | 6 |

**Table 8** Raw IS intensities and individual coefficients of variation (n=5) used for calibration curves - for plasma (upper part of the table) and urine samples (bottom part of the table).

| SM 30:1;O2 | | | SHexCer 30:1;O2 | | |
|---|---|---|---|---|---|
| c [nmol/mL] | Median of Intensity | CV[%] | c [pmol/mL] | Median of Intensity | CV [%] |
| 8.66 | 2.80E+05 | 6 | 20 | 2.23E+04 | 5 |
| 17.32 | 6.35E+05 | 17 | 40 | 6.67E+04 | 15 |
| 25.99 | 8.90E+05 | 8 | 60 | 9.26E+04 | 3 |
| 34.65 | 1.09E+06 | 11 | 80 | 1.17E+05 | 5 |
| 43.31 | 1.50E+06 | 5 | 100 | 1.56E+05 | 8 |
| 51.97 | 1.67E+06 | 6 | 120 | 1.95E+05 | 4 |
| 69.3 | 2.28E+06 | 6 | 160 | 2.66E+05 | 7 |
| 86.62 | 3.11E+06 | 10 | 200 | 3.44E+05 | 9 |

| SHexCer 30:1;O2 | | | Taurocholic acid-d4 | | |
|---|---|---|---|---|---|
| c [pmol/mL] | Median of Intensity | CV [%] | c [nmol/mL] | Median of Intensity | CV [%] |
| 2.25 | 1.13E+04 | 13 | 0.12 | 1.65E+04 | 10 |
| 4.50 | 4.29E+04 | 20 | 0.25 | 5.84E+04 | 7 |
| 8.99 | 6.93E+04 | 10 | 0.62 | 9.73E+04 | 11 |
| 22.49 | 1.38E+05 | 11 | 1.23 | 2.38E+05 | 5 |
| 44.97 | 3.35E+05 | 10 | 2.46 | 5.80E+05 | 6 |
| 89.94 | 8.95E+05 | 4 | 6.15 | 1.22E+06 | 5 |
| 224.85 | 1.53E+06 | 6 | 12.30 | 2.27E+06 | 5 |
| 449.71 | 3.45E+06 | 9 | 24.61 | 5.77E+06 | 7 |

**Table 9** Individual recovery rates calculated based on the raw data in Supplementary Table 5 together with average values and coefficients of variation obtained for the extraction protocol of pooled plasma sample for applied internal standards, where abbreviation LL means low concentration level, ML means medium concentration level, and HL means high concentration level.

| Level | Sample No. | Recovery rate [%] | |
|---|---|---|---|
| | | SM 30:1;O2 | SHexCer 30:1;O2 |
| Low level | LL1 | 105 | 130 |
| | LL2 | 102 | 121 |
| | LL3 | 99 | 121 |
| | LL4 | 90 | 101 |
| | LL5 | 91 | 125 |
| | LL6 | 100 | 107 |
| | Average | 98 | 118 |
| | CV [%] | 6 | 9 |
| Medium level | ML1 | 100 | 124 |
| | ML2 | 94 | 116 |
| | ML3 | 115 | 114 |
| | ML4 | 85 | 96 |
| | ML5 | 105 | 96 |
| | ML6 | 95 | 109 |
| | Average | 99 | 109 |
| | CV [%] | 10 | 10 |
| High level | HL1 | 87 | 111 |
| | HL2 | 115 | 116 |
| | HL3 | 93 | 114 |
| | HL4 | 90 | 119 |
| | HL5 | 103 | 103 |
| | HL6 | 92 | 94 |
| | Average | 97 | 109 |
| | CV [%] | 10 | 8 |

**Table 10** Individual recovery rates calculated based on the raw data in Supplementary Table 7 together with average values and coefficients of variation obtained for the extraction protocol of the pooled urine sample for applied internal standards, where abbreviation LL means low concentration level, ML means medium concentration level, and HL means high concentration level.

| Level | Sample No. | Recovery rate [%] | |
|---|---|---|---|
| | | **SHexCer 30:1;O2** | **Taurocholic acid-d4** |
| Low level | LL1 | 85 | 119 |
| | LL2 | 84 | 106 |
| | LL3 | 98 | 83 |
| | Average | 89 | 102 |
| | CV [%] | 6 | 15 |
| Medium level | ML1 | 105 | 101 |
| | ML2 | 83 | 98 |
| | ML3 | 93 | 120 |
| | Average | 94 | 106 |
| | CV [%] | 9 | 10 |
| High level | HL1 | 96 | 116 |
| | HL2 | 111 | 123 |
| | HL3 | 93 | 91 |
| | Average | 100 | 110 |
| | CV [%] | 8 | 14 |

**Table 11** Coefficients of variation representing within (n=3) and between (n=6) days precision for internal standards applied for plasma measurements (upper part of the table) and urine measurements (bottom part of the table). Individual verage intensities and CV were calculated from the absolute intensities of Supplementary Tables 5 (plasma) and 7 (urine). The abbreviation LL means low concentration level, ML means medium concentration level and HL means high concentration level.

| Type | Level | SM 30:1;O2 | | SHexCer 30:1;O2 | |
|---|---|---|---|---|---|
| | | **Average Intensity** | **CV [%]** | **Average Intensity** | **CV [%]** |
| Within day presicion | LL | 7.81E+05 | 4 | 9.04E+04 | 9 |
| | ML | 1.04E+06 | 4 | 1.14E+05 | 1 |
| | HL | 1.83E+06 | 9 | 2.26E+05 | 2 |
| Between day precision | LL | 7.95E+05 | 4 | 9.66E+04 | 9 |
| | ML | 1.03E+06 | 7 | 1.25E+05 | 9 |
| | HL | 1.80E+06 | 7 | 2.41E+05 | 4 |

| Type | Level | SM 30:1;O2 | | SHexCer 30:1;O2 | |
|---|---|---|---|---|---|
| | | **Average Intensity** | **CV [%]** | **Average Intensity** | **CV [%]** |
| Within day presicion | LL | 2.86E+04 | 15 | 1.59E+04 | 14.6 |
| | HL | 5.03E+05 | 7 | 3.05E+05 | 2.3 |
| Between day precision | LL | 2.56E+04 | 22 | 1.59E+04 | 22.0 |
| | HL | 4.34E+05 | 17 | 2.72E+05 | 15.1 |

**Table 12** Matrix effect of internal standards used for plasma measurements. The matrix effect is expressed by the coefficient of variation of average signal response within 6 different plasma samples (individual raw intensities are shown in Supplementary Table 6) at three concentration levels, where the abbreviation LL means low concentration level, ML means medium concentration level, and HL means high concentration level.

| **Level** | **SM 30:1;O2** | | **SHexCer 30:1;O2** | |
|---|---|---|---|---|
| | **Average Intensity** | **CV [%]** | **Average Intensity** | **CV [%]** |
| LL | 9.33E+05 | 12 | 8.40E+04 | 9 |
| ML | 1.16E+06 | 14 | 1.08E+05 | 13 |
| HL | 1.91E+06 | 16 | 1.95E+05 | 13 |

**Table 13** The upper part of the table shows the freeze/thaw stability of internal standards at medium concentration level applied for plasma measurements. Freeze/thaw stability is expressed by the coefficient of variation of the average signal response for 6 individual plasma samples after 3 freeze/thaw cycles (individual raw intensities are shown in Supplementary Table 6). The bottom part of the table shows freeze/thaw stability of internal standards applied for urine measurements. The freeze/thaw stability is expressed by the coefficient of variation (n=3) of average signal response for the pooled sample at a low (LL) and high (HL) concentration levels after one freeze/thaw cycle (individual absolute intensities of LL4 – LL6 and HL4 – HL6 before and after freeze/thaw cycle are shown in Supplementary Table 8).

| Sample No | SM 30:1;O2 | | SHexCer 30:1;O2 | |
|---|---|---|---|---|
| | Average Intensity | CV [%] | Average Intensity | CV [%] |
| M_31 | 1.04E+06 | 18 | 1.03E+05 | 18 |
| M_43 | 1.03E+06 | 17 | 1.03E+05 | 25 |
| M_96 | 9.65E+05 | 11 | 9.41E+04 | 14 |
| M_97 | 8.93E+05 | 10 | 8.66E+04 | 16 |
| M_101 | 1.17E+06 | 12 | 8.35E+04 | 23 |

| Level | Taurocholic acid-d4 | | SHexCer 30:1;O2 | |
|---|---|---|---|---|
| | Average Intensity | CV [%] | Average Intensity | CV [%] |
| LL | 1.39E+04 | 27 | 2.42E+04 | 24.4 |
| HL | 2.29E+05 | 14.5 | 3.44E+05 | 9.8 |
